# Data-driven trajectories of atrophy explain clinical heterogeneity across Lewy body diseases

**DOI:** 10.64898/2026.01.29.26344990

**Authors:** Ajay Konuri, Gonzalo Castro Leal, Niloufar Zebarjadi, Annegret Habich, Nicolás Castellanos-Perilla, María Camila Gonzalez, John-Paul Taylor, Michael Firbank, Daniel Alcolea, Alexandre Bejanin, Kurt Segers, Florence Benoit, Ahmet Turan Isik, Bedia Samanci, Consuelo Cháfer-Pericás, Richard Wade-Martins, Michele T M Hu, Rohan Bhome, Ivelina Dobreva, Zuzana Walker, Dag Aarsland, Eric Westman, Daniel Ferreira, Glenda Halliday, Simon JG Lewis, Rimona S Weil, Ramon Landin-Romero, Christian Lambert, Neil P. Oxtoby, Elie Matar

## Abstract

**Background:** Lewy body diseases (LBD) collectively share α-synuclein Lewy pathology, yet present wide clinical heterogeneity, with overlapping motor and non-motor features and progression patterns that challenge traditional diagnostic boundaries.

**Methods:** To resolve this spatiotemporal heterogeneity at the biological level, we applied a data-driven atrophy progression framework to MRI data from 833 individuals across Parkinson’s disease (PD), dementia with Lewy bodies (DLB), and prodromal idiopathic REM sleep behaviour disorder (iRBD) using the Subtype and Stage Inference (SuStaIn) algorithm.

**Findings:** Four transdiagnostic subtypes (A: Early cortico-limbic/late basal ganglia, B: Early basal ganglia/late limbic, C: Early temporo-limbic/late basal ganglia, and D: Early basal ganglia–cingulate/late cortex) emerged, each defined by a distinct spatiotemporal progression of atrophy that explained cognitive, motor, and psychiatric variability. An early cortico-limbic/late basal ganglia subtype represented a dementia-prone subtype across clinical diagnoses, with limbic involvement associating with the emergence of visual hallucinations.

**Interpretation:** These biologically relevant spatiotemporal atrophy subtypes provide an interpretable stratification of patients with LBD, with potential to refine prognosis, improve clinical trial stratification, and guide precision therapeutic approaches.

**Funding:** This work was made possible by an Ignition grant from the University of Sydney and University College London (Global Engagement Fund).

**Research in context:** *Evidence before this study:* Parkinson’s disease (PD), dementia with Lewy bodies (DLB), and isolated REM sleep behaviour disorder (iRBD), sit on a Lewy body disease spectrum with overlapping clinical features but marked heterogeneity in symptom profile, timing, and progression. Neuroimaging and neuropathological studies report diverse patterns of neurodegeneration within each syndrome, with partial overlap across the broader spectrum. Recent, large-scale subtyping work suggests at least two spatiotemporal neurodegeneration trajectories. However, *in vivo* imaging subtyping which has particularly strong translational relevance, has largely focused on a single syndrome or has included limited representation of the full Lewy body disease continuum.

*Added value of this study:* This study includes a large, balanced cohort spanning PD, DLB and prodromal iRBD with detailed clinical and neuropsychological phenotyping. Using MRI-derived atrophy, we identified four transdiagnostic subtypes that capture clinically meaningful variability beyond syndromic diagnosis. We found that stage, an indicator of accumulated atrophy better addresses the between-syndromic differences in disease duration. We also show that certain subtypes with early limbic degeneration are associated with visual hallucinations, and are linked to amygdalar atrophy, and that subtype plus stage improves prediction of hallucinations beyond amygdalar volume alone. Longitudinal analysis further identified a subset of patients with PD with earlier cortico-limbic degeneration as being at higher risk of cognitive impairment.

*Implications of all the available evidence:* The current field of LBD is moving towards a biologically grounded stratification and staging criteria that better reflects clinical heterogeneity. These transdiagnostic subtypes alongside stage, provide complementary information that can inform emerging LBD staging frameworks and enable more targeted clinical trial design through improved stratification and risk enrichment.

## Introduction

Lewy body diseases (LBD) are the second most common cause of neurodegeneration^1^ and include the canonical clinical syndromes of Parkinson’s disease (PD), dementia with Lewy bodies (DLB), and isolated rapid eye movement (REM) sleep behaviour disorder (iRBD). These syndromes are unified by the pathological hallmark of α-synuclein Lewy pathology in neurons. PD is diagnosed by the presence of bradykinesia, rigidity, and tremor,^2^ whereas DLB is diagnosed by functionally limiting cognitive impairment accompanied by at least two core clinical features of visual hallucinations, cognitive fluctuations, REM sleep behaviour disorder (RBD), and parkinsonism.^3^ iRBD, characterised by dream enactment behaviours and polysomnographic loss of REM atonia, is well established as a prodromal disorder, with more than 75% of patients progressing to an overt α-synucleinopathy (mostly PD and DLB).^4^ Despite these apparently discrete clinical syndromes, patients ultimately converge on overlapping motor, cognitive, psychiatric, sleep, and autonomic features.^5^ This overlap has motivated proposals to unify LBD within revised diagnostic criteria and biological staging frameworks, requiring a reappraisal of how to best delineate different pathways of neurodegeneration that may give rise to such different clinical syndromes (reviewed in ^6^).

Historically, distinctions between PD and DLB have relied on the temporal separation of motor and cognitive symptoms, most notably the ‘one-year rule’ that distinguishes Parkinson’s disease dementia (PDD) from DLB as dementia arising more than one year after parkinsonism. Although pragmatic, such heuristics do not reflect biological boundaries, and there is striking heterogeneity within each syndrome. In PD, some patients remain motor-predominant for decades, whereas others manifest early and significant non-motor symptoms, including cognitive decline, RBD, and autonomic dysfunction. These differences are captured in both clinical and data-driven clustering studies that distinguish ‘mild motor predominant’ and ‘diffuse malignant’ subtypes, with the latter more closely resembling DLB.^7^ Within DLB, subsets present with early psychiatric or visuospatial deficits, whereas others follow a motor-predominant trajectory.^3,8^ Even iRBD exhibits variable motor, cognitive, and autonomic features that may foreshadow divergent outcomes.^9^ Understanding and predicting these trajectories, particularly in iRBD, remains a critical challenge for prognosis and for enriching trials of disease-modifying interventions.

Until recently, biologically driven efforts to subtype and stage all LBD together have been largely restricted to post-mortem cohorts.^6^ Pathological subtyping and staging systems^8,10–12^ and more recent data-driven frameworks^13,14^ emphasise at least three spatiotemporal patterns of spread of Lewy pathology. Previous efforts have demonstrated that individuals with PD and DLB can be distributed (albeit unevenly) across all pathological subtypes, implying substantial variability in pathological progression within each syndrome and only partial correlation with symptom timing.^12–14^ Frameworks based only on neuropathology are inherently limited by reliance on end-stage tissue and late-life clinical information, and clinicopathological studies show that the relationship between pathology and clinical presentation is heterogeneous. Lewy pathology is not always aligned with clinically defined PD, with some individuals developing typical symptoms despite little or no α-synuclein pathology, whereas others with incidental Lewy body pathology at autopsy never manifest symptoms, underscoring the need for additional biomarkers of progression. Cell loss, reflected in part by atrophy, appears more linked to symptoms in LBD ^6^, with vulnerability dictated by several cellular factors (e.g., mitochondrial dysfunction, altered calcium handling, impaired proteostasis, and sustained oxidative stress).^15,16^

Structural neuroimaging provides a non-invasive and widely accessible *in vivo* measure of neurodegeneration and has revealed both distinct and overlapping atrophy patterns across LBD. The earliest structural changes in PD are typically observed in the striatum,^17^ hippocampus,^18^ and the limbic system.^19,20^ With advancing disease, atrophy extends to the thalamus and neocortex, often coinciding with the onset of cognitive impairment.^21^ In DLB, atrophy consistently affects the neocortex, insula, putamen, thalamus, and the limbic system^22,23^ and is more severe than PDD^24,25^ but less than Alzheimer’s disease (AD).^26,27^ Although less pronounced, early-stage atrophy is also seen in iRBD, particularly within the basal ganglia, cingulate^28,29^ and limbic structures.^30^ Within this broader pattern of neurodegeneration, widespread cortical, limbic and thalamic atrophy has been associated with visual hallucinations.^31^ However, recent structural and functional imaging evidence implicates the amygdala as a central convergence hub, integrating visual and limbic processes,^32,33^ with voxel-level mapping highlighting it as a dominant locus of visual dysfunction relative to adjacent limbic structures.^34^ Despite these insights, inconsistencies exist across studies,^27,35^ in part because conventional group-level analyses are vulnerable to the significant clinical heterogeneity in these cohorts. In particular, variability in subtype representation and disease stage or severity complicates the interpretation of imaging findings and contributes to inconsistency across studies. This has limited the identification of a reproducible imaging biomarker for core clinical features such as visual hallucinations.

In view of this, recent work has shifted towards using objective biological measures for data-driven staging frameworks that explicitly model heterogeneity and disease progression.^6^ Subtype and Stage Inference (SuStaIn) is an unsupervised machine learning technique that stratifies patients into subtypes and stages based on phenotypic and temporal disease progression.^36,37^ Several recent neuroimaging studies have demonstrated the utility of SuStaIn to predict subtypes and their progression across various conditions including AD,^38,39^ frontotemporal dementia^36^ and multiple sclerosis^40^. So far, *in vivo* applications of SuStaIn in PD cohorts have suggested two clinical^41^ and three structural imaging subtypes in PD.^42,43^ To date, application of SuStaIn in DLB has been limited to a single small PET imaging study identifying two progression patterns^44^, and a study in iRBD incorporating relatively few participants with established PD or DLB.^45^ A separate memory clinic study applied SuStaIn as a transdiagnostic effort to subtype AD and mild cognitive impairment (MCI), with limited representation of LBD.^46^ Across these cohorts, the over-representation of a single clinical diagnosis and under-representation of DLB may have constrained the ability to capture true biological heterogeneity across LBD and limited the clinical utility of the models.

Recognising the need for approaches that target biological neurodegeneration across all LBD, we applied SuStaIn to a large multi-cohort dataset spanning iRBD, PD, and DLB. We hypothesised that joint consideration of subtype and stage of neurodegeneration across these diagnoses would provide a biologically grounded framework for explaining the heterogeneous emergence and progression patterns of cognitive, motor, neuropsychiatric, and RBD symptoms. We identified four robust atrophy trajectories that cut across conventional diagnostic categories and were associated with distinct clinical profiles, including an aggressive dementia-predominant subtype. Visual hallucinations were most prevalent in subtypes with early limbic involvement. In a subset of individuals with iRBD, baseline subtype assignment was related to the subsequent phenoconversion pathway to PD or DLB. Finally, validation analyses using longitudinal clinical data available in individuals with PD revealed that baseline subtype correlated with risk of cognitive impairment in PD, underscoring the clinical relevance of these trajectories.

## Methods

This study involved analysis of a multi-cohort international dataset of 1,373 participants who had undergone T1-weighted MRI imaging. This included 506 healthy controls (HC), 294 clinically diagnosed individuals with DLB, 129 with iRBD, and 444 patients with PD, of whom 33 had PDD. MCI with Lewy bodies (MCI-LB) were not included in this study due to limited available data. Biological sex was self-reported by the participants. Data came from seven multicentre cohorts across 12 recruitment centres including the E-DLB consortium (n=400), National Alzheimer’s Coordinating Centre (NACC) (n=32), Oxford Parkinson’s Disease Centre Discovery Cohort (OPDC) (n=221), Parkinson’s Disease Research Clinic (University of Sydney & Macquarie University) (n=214), Parkinson’s Progression Markers Initiative (PPMI) (n=339), Vision in Parkinson’s (VIP) (n=56) and Quantitative MRI for Anatomical Phenotyping in Parkinson’s Disease (qMAP-PD) (n=111) (Supplementary Table S1). Data used in the preparation of this article were obtained on 2023-10-26 from the Parkinson’s Progression Markers Initiative (PPMI) database (https://www.ppmi-info.org/access-data-specimens/download-data), RRID:SCR_006431. For up-to-date information on the study, visit http://www.ppmi-info.org.

### Data Preparation, Feature Selection and Outlier Detection

Cross-sectional structural T1 scans were pre-processed using FreeSurfer version 7.1.1, except for the E-DLB cohort, which was processed using FreeSurfer version 7.3 (http://surfer.nmr.mgh.harvard.edu). Cortical and subcortical volumes for all centres were derived using the Desikan-Killiany (DK) atlas. The estimated total intracranial volume (TIV) was regressed out to normalise regional brain volumes.

Feature selection was based on a hybrid data-driven and evidence-based approach as per an *a priori* analysis plan. Principal component analysis (PCA) was first applied to regional volumes, and regions with the strongest PC1 loadings were retained. PC1 explained 37.8% of the total variance, with the strongest loadings observed in frontal and posterior association cortices (Supplementary Table S2). These were cross-checked against prior published structural imaging literature in PD and DLB to ensure anatomical relevance to LBD. Final ROIs are detailed in Supplementary Table S2 and on the y-axis of Figure 1, and Supplementary Figures S6–S7.

**Figure 1:**
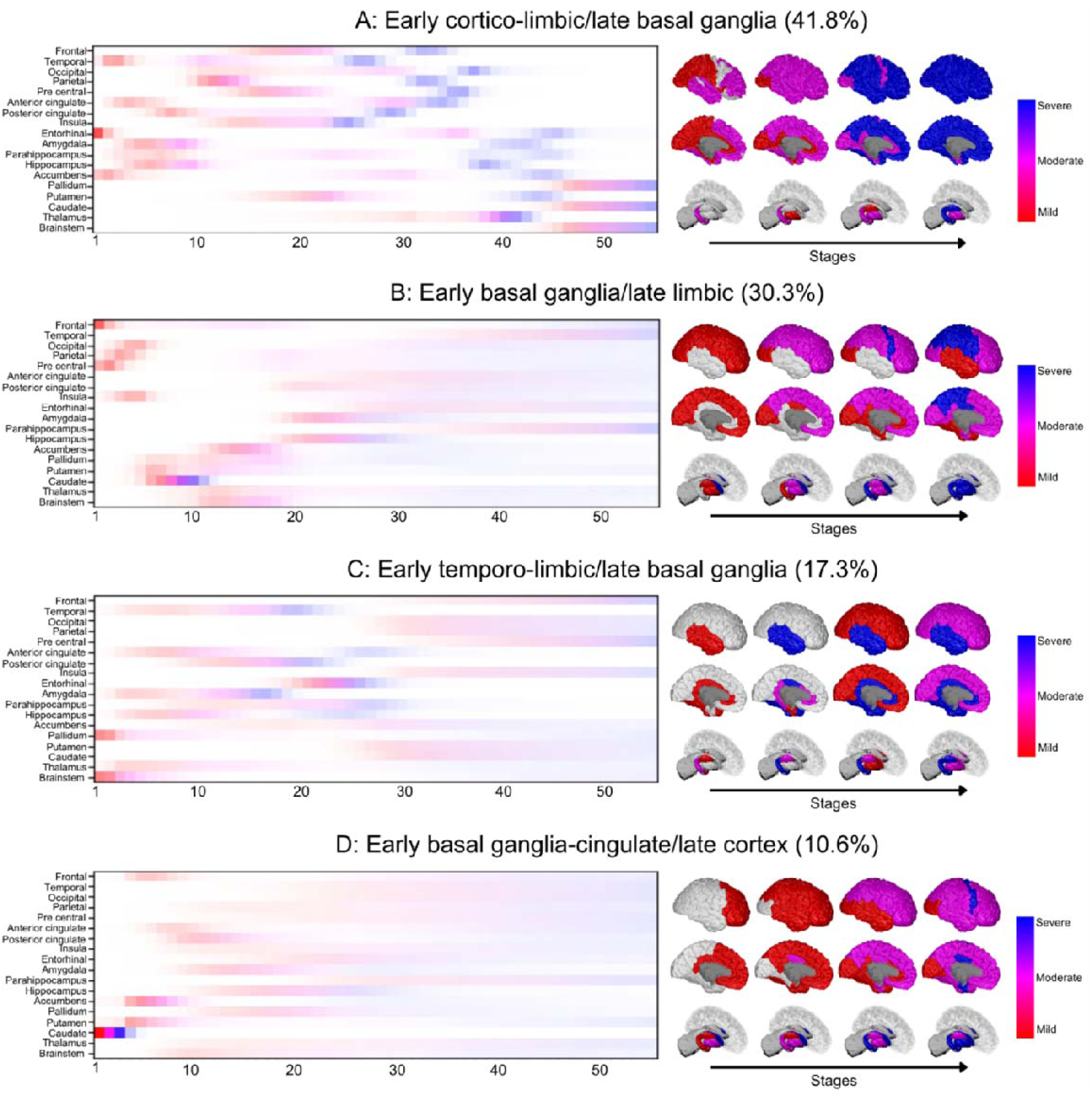
Spatiotemporal atrophy signatures of LBD subtypes. Data-driven subtypes show distinct sequences of regional degeneration across all LBD. Positional variance diagrams show atrophy by SuStaIn stage (x-axis) for cortical and subcortical regions (y-axis). Percentages indicate the proportion of 710 individuals with LBD in each subtype after excluding 123 Stage 0 individuals. Subtypes are defined based on the ordering of severe atrophy events, capturing the most discriminative patterns of neurodegeneration. **A:** Early cortico-limbic/late basal ganglia (41.8%). Atrophy progresses in a rostro-caudal axis with atrophy initiating in neocortex before progressing to subcortical structures. **B:** Early basal ganglia/late limbic (30.3%). Basal ganglia are affected earlier than rest of the brain, with relative preservation of the limbic structures. **C:** Early temporo-limbic/late basal ganglia (17.3%). Temporal and limbic cortices lead, with basal ganglia changes appearing later. **D:** Early basal ganglia–cingulate/late cortex (10.6%). Diffuse mild degeneration across the brain with severe atrophy seen in basal ganglia. Colours denote atrophy level and transparency indicates the probability of z-scores, Red: Mild atrophy (z > 0.5), Magenta: Moderate atrophy (z > 1.0), Blue: Severe atrophy (z > 2.0).

In controls, volumetric outliers per DK atlas ROI were defined as values exceeding three standard deviations from the group mean. To avoid censoring meaningful deviations based on pathology in the disease groups (DLB, iRBD and PD), a multivariate approach to outlier removal was applied based on the Mahalanobis distance^47–49^ between each subject’s vector of regional brain volumes and the group centroid estimated with a robust minimum covariance determinant estimator based on the selected ROIs. The threshold for Mahalanobis distance was determined using a chi-square distribution threshold corresponding to the 99^th^ percentile. In total, 88 scans were excluded (19 DLB, 8 PD, 7 iRBD, and 54 HC).

### Subtype and Stage Inference

We applied the SuStaIn algorithm, implemented via the pySuStaIn package^37^ (see pySuStaIn repository: https://github.com/ucl-pond/pySuStaIn), to simultaneously uncover data-driven subtypes of spatiotemporal atrophy from regional brain volumes. Unlike conventional clustering algorithms, which group individuals based on static features of similarity, SuStaIn jointly models disease progression and heterogeneity by inferring subtype-specific sequences of biomarker change alongside an individual’s stage within that sequence. Disease progression is modelled as a series of piecewise-linear z-score transitions between selected thresholds (z=0.5, 1, 2), representing increasing levels of atrophy. Each cluster/subtype captures a distinct spatiotemporal sequence of regional involvement, while the SuStaIn stage represents cumulative progression along that subtype-specific path. We ran SuStaIn using 25 start points and 1,000,000 MCMC iterations. Individuals were assigned to subtypes based on maximum likelihood estimates.

To capture transdiagnostic trajectories of clinical LBD, we first trained a combined model including all three clinical groups (PD, DLB, and iRBD) projecting them onto a unified disease continuum. We then trained two additional syndrome-specific models, one restricted to PD and another to DLB, to benchmark against prior subtype solutions of PD and to delineate the structural subtypes in DLB. Using 18 regional features and three severity levels, each subtype trajectory comprised 54 discrete stages (18 features × 3 z scores).

Model complexity was assessed using out-of-sample predictions under 10-fold cross-validation. For each fold, SuStaIn was trained on 90% of the dataset, and predictive performance was assessed on the held out 10%. Model fit was quantified using both the Cross-Validation Information Criterion (CVIC) and out-of-sample log-likelihood. These metrics improved with increasing subtype number but showed diminishing returns beyond four subtypes in the combined model and three subtypes in the disease-specific models, supporting selection of a parsimonious solution (Supplementary Figure S1). Subtype assignment probabilities increased with advancing disease stage (Supplementary Figure S2), consistent with previous SuStaIn studies using imaging biomarkers,^41,50^ displaying greater confidence in classification later in the disease. Individuals without classifiable regional atrophy (grouped as Stage 0) were excluded from clinical phenotyping.

### Clinical Assessments

Cognition was assessed using the Mini-Mental State Examination (MMSE) and Montreal Cognitive Assessment (MoCA), and motor impairment with the Movement Disorders Society Unified Parkinson’s Disease Rating Scale: Part III Motor Examination (MDS-UPDRS-III). Non-motor symptoms included REM Sleep Behaviour Disorder Screening Questionnaire (RBDSQ), Hospital Anxiety and Depression Scale (HADS-A and HADS-D), and Trail Making Task Part B (TMT-B) for executive function. Autonomic symptom burden across symptoms including pain, urinary dysfunction, constipation, light-headedness, and fatigue was examined across Parkinson’s disease (PD) subtypes using relevant items from the MDS-UPDRS Part I (non-motor experiences of daily living).

Presence or absence of core clinical features was recorded for PD and DLB cohorts, including visual hallucinations (281 PD, 271 DLB) and RBD (141 PD, 230 DLB). Participants with iRBD were excluded from these analyses as RBD is universal in this group and visual hallucinations typically emerge only after phenoconversion. Longitudinal iRBD conversion data were available for 41 participants from Sydney.

### α-synuclein seed amplification assay (SAA)

α-synuclein SAA data were available for a subset of participants in the PPMI cohort (n=131), collected at the same MRI visit. α-synuclein seeding activity was assessed using Amprion SAA. Full biomarker acquisitions are available at https://ida.loni.usc.edu/pages/access/.

### Dopamine Transporter Imaging (DaT-SPECT)

DaT-SPECT imaging using [¹²³I]-FP-CIT SPECT was available for participants with early-stage PD in the PPMI cohort. Longitudinal striatal binding ratios (SBR) were calculated using the procedures outlined in the PPMI imaging technical operations manual (http://ppmi-info.org/).

### Statistical Analysis

Optimal subtype, stage, and probability assignments were derived using *pySuStaIn*’s built-in assignment algorithm.^37^ Individuals with LBD having SuStaIn stages ≥ 1 were compared across classified subtypes, along with an additional unclassified group. For statistical modelling, SuStaIn stage was treated as a continuous variable to preserve statistical power and avoid over-parameterisation across a large number of stages consistent with prior studies.^42^ To capture potential disease-specific nuances, final subtype distributions of the combined LBD model were further examined within each clinical diagnosis.

Demographic variables (age, sex, education, disease duration from diagnosis) and clinical measures were compared across subtypes using non-parametric permutation tests (10,000 permutations) with *FDR* correction for multiple comparisons. Categorical variables were assessed using χ² tests.

The presence of visual hallucinations and RBD across SuStaIn stages in PD and DLB was investigated using multivariable logistic regression, adjusting for age and sex, with stage, diagnosis, and their interaction as predictors. Associations between SuStaIn stage and continuous clinical variables were examined using linear regression, adjusting for age and sex (clinical variable ∼ age + sex + Subtype × Stage). Stage-based analysis was performed using subtype-specific transition points derived from the SuStaIn model. Limbic involvement was defined based on the stage at which limbic structures became moderately affected (z>1) within each subtype progression pattern. Patients were subsequently classified as pre-limbic (stage below the subtype-specific threshold) or post-limbic (stage at or beyond the threshold). Logistic regression models were used to evaluate the association between post-limbic status and visual hallucinations, adjusting for subtype, age, and sex. To examine the incremental predictive utility of SuStaIn-derived parameters over structural volumetry for visual hallucinations, three logistic regression models were fit in patients with PD, incorporating amygdala z-score, subtype assignment, and their combination with stage as predictors. Non-nested models were compared against an intercept-only null model and nested model discrimination was compared using the DeLong method.

RBD diagnosis was confirmed using overnight polysomnography in the iRBD cohort and in a subset of PD and DLB participants and was otherwise assessed using the REM Sleep Behaviour Disorder Screening Questionnaire (RBDSQ). A binomial test was used to determine whether the observed proportion of iRBD converters in a given subtype significantly exceeded the expected proportion under a uniform distribution across the classified subtypes. Fisher’s exact test was used to compare subtype distributions directly between PD and DLB converters.

Longitudinal analyses of cognitive, motor, and RBD symptom progression were done using the available data from PPMI and Sydney cohorts (n=137). For each clinical measure, patients with the following number of follow-ups were available: MoCA (n=135): ( ≥3: n=94; 2: n=16; 1: n=25), MDS-UPDRS-III (n=131): (≥3: n=93; 2: n=16; 1: n=22); RBDSQ (n=35) (≥3: n=30; 2: n=2; 1: n=3). Linear mixed effects regression models were fitted for MoCA, MDS-UPDRS-III, and RBDSQ with fixed effects for time (months since baseline), age, sex, SuStaIn stage, subtype, and a time-by-subtype interaction. A subject-specific random intercept was included to account for repeated within-subject measurements. RBDSQ analyses were restricted to participants with evidence of RBD at baseline (RBDSQ ≥ 5).

Risk of developing mild cognitive impairment (MCI) within PD was assessed using log-rank tests and Cox proportional hazards models based on the same longitudinal MoCA data, with adjustment for age, sex, and SuStaIn stage. Model fit was assessed using the log-likelihood ratio test. Kaplan–Meier curves were generated for visualisation, and hazard ratios with 95% confidence intervals were reported.

Across all clinical regression and longitudinal models, SuStaIn model stage was included as a covariate to account for progression along each participant’s assigned subtype-specific trajectories, unless otherwise stated. Model assumptions were assessed for all analyses. For linear and linear mixed-effects models, residual diagnostics were used to evaluate model fit. Linear mixed-effects models included subject-specific random intercepts to account for repeated measurements. Missing data were assumed to be missing at random, and this assumption was examined by evaluating patterns of missingness across subtypes and testing associations with observed baseline variables. For Cox proportional hazards models, the proportional hazards assumption was assessed using Schoenfeld residuals. Statistical significance was defined at p*_FDR_*<0.05 with false discovery rate (*FDR*) correction for multiple comparisons applied separately within each predefined analysis family corresponding to a specific research question or hypothesis; uncorrected p values are explicitly labelled where reported.

### Ethics approval and consent to participate

All patients gave written informed consent according to the Declaration of Helsinki, and ethics approval was obtained from the local institutional boards. This study was undertaken as part of the Transdiagnostic Subtyping and Classification Effort in Neurodegenerative Disease (TranSCEND) initiative (scripts and code made available at https://ucl-usyd-transcend.github.io/). Analysis of this data was approved by the Sydney University Human Research Ethics Committee (HREC No. 2013/HE000945), UCL Research Ethics Committee under application 8019/005 and the UCL Department of Computer Science Research Ethics Committee under application UCL-CSREC-209-B.

## Results

We analysed T1-weighted MRI data from 1,285 participants across 12 centres. The cohort included 833 patients with clinically diagnosed LBD (PD, n=436; DLB, n=275, and iRBD, n=122), and 452 HC. α-synuclein SAA data were available for a subset of patients with PD (n=131) and showed a high positivity rate (88.5%). Detailed demographics by clinical diagnoses are provided in Supplementary Table S1. Patients with DLB were significantly older than those with PD, iRBD, and HC (all p*_FDR_*<0.001). Disease groups had a higher proportion of males compared to HC (p*_FDR_*<0.001), consistent with known sex differences in these populations.^51^

### Subtypes of spatiotemporal atrophy patterns in LBD

Application of SuStaIn to the combined LBD population with 10-fold cross-validation (Supplementary Figure S1) indicated that a four-subtype solution provided the best fit with disease progression stratified across 54 distinct stages (Figure 1). Among 833 participants with LBD, 123 (14.7%) were assigned to Stage 0 (‘S0’) with no detectable atrophy and were excluded from subsequent analyses (Demographics and clinical composition of S0 are mentioned in Table 1, and Supplementary Table S3).

**Table 1:** Summary of demographics and clinical features by subtypes. Displayed values indicate median [95% CI] of demographic and clinical data across the four classified subtypes and the unclassified group. Disease durations (years) are reported for each diagnosis. Stage is mean ± SD. Pairwise post-hoc comparisons (FDR corrected, p<0.05) are denoted as: * indicates significant differences between classified subtypes; ^ indicates differences between the unclassified and classified subtypes. Letter markers denote pairwise comparisons among classified subtypes: a (A vs B), b (A vs C), c (A vs D), d (B vs C), e (B vs D). HADS (Hospital Anxiety and Depression scale), MDS-UPDRS–III (Movement Disorder Society Unified Parkinson’s Disease Rating Scale Section III, MMSE (Mini Mental State Examination), MoCA (Montreal Cognitive Examination).

| Variables | Stage 0 (n=123) | Early cortico-limbic/late basal<br>ganglia (A, n=297) | Early basal<br>ganglia/late limbic (B, n=215) | Early temporo-limbic/late basal<br>ganglia (C, n=123) | Early basal ganglia-<br>cingulate/late cortex<br>(D, n=75) |
| --- | --- | --- | --- | --- | --- |
| Age* | 68 [66, 69]<br>(n=123) | 71 [69, 73]<sup>a,b</sup><br>(n=297) | 68 [66, 69]<sup>a</sup><br>(n=215) | 67 [66, 70]<sup>b</sup><br>(n=123) | 70 [68, 72]<br>(n=75) |
| Sex (M:F) | 89:34 | 198:99<sup>c</sup> | 134:81<sup>e</sup> | 97:26 | 66:9<sup>c,e</sup> |
| Education<sup>^*</sup> | 15 [14, 15]<sup>^</sup><br>(n=105) | 12 [12, 13]<sup>^,a,b,d</sup><br>(n=287) | 15 [15, 16]<sup>a</sup><br>(n=163) | 15 [15, 15]<sup>b</sup><br>(n=114) | 13 [10, 15]<sup>d</sup><br>(n=65) |
| Disease Duration - DLB<br>(years)* | 1.8 [0.2, 4.0]<br>(n=18) | 1.2 [0.7, 1.7]<br>(n=86) | 0.7 [0, 0.9]<sup>e</sup><br>(n=20) | 1.8 [1.1, 3.0]<br>(n=21) | 3.9 [1.2, 5.0]<sup>e</sup><br>(n=11) |
| Disease Duration - PD<br>(years)*<sup>^</sup> | 1 [0.7, 1.7]<sup>^</sup><br>(n=66) | 1.8 [1.4, 2.3]<sup>^,c</sup><br>(n=88) | 1.6 [1.4, 2.0]<sup>d,e</sup><br>(n=165) | 2.7 [2.0, 4.1]<sup>^,d</sup><br>(n=36) | 4.3 [2.8, 6.3]<sup>^,c,e</sup><br>(n=25) |
| Disease Duration - iRBD<br>(years) | 0.6 [0.5, 1.7]<br>(n=18) | 1.9 [1, 2.5]<br>(n=34) | 0.83 [0.41, 1.33]<br>(n=6) | 1.3 [0.9, 2.4]<br>(n=36) | 1.1 [0.6, 1.3]<br>(n=24) |
| DLB - n (%) | 33 (26.8%) | 160 (53.9%) | 31 (14.4%) | 31 (25.2%) | 20 (26.7%) |
| PD - n (%) | 75 (61.0%) | 101 (34.0%) | 177 (82.3%) | 56 (45.5%) | 27 (36.0%) |
| iRBD - n (%) | 15 (12.2%) | 36 (12.1%) | 7 (3.3%) | 36 (29.3%) | 28 (37.3%) |
| **Questionnaires** |  |  |  |  |  |
| HADS Anxiety | 4 [3, 6]<br>(n=35) | 4 [3, 5]<br>(n=78) | 3 [2, 4]<br>(n=41) | 4 [2, 6]<br>(n=53) | 2.5 [1, 4]<br>(n=24) |
| HADS Depression | 4 [2, 5]<br>(n=35) | 3 [2, 4]<br>(n=78) | 3 [2, 4]<br>(n=41) | 4 [2, 5]<br>(n=53) | 2 [0, 4]<br>(n=24) |
| **Cognitive Measures** |  |  |  |  |  |
| MMSE*<sup>^</sup> | 27 [26, 29]<sup>^</sup><br>(n=62) | 27 [26, 27]<sup>a</sup><br>(n=209) | 28 [28, 29]<sup>^,a,d,e</sup><br>(n=97) | 26 [24, 26]<sup>d</sup><br>(n=87) | 27 [26, 28]<sup>e</sup><br>(n=34) |
| MoCA*<sup>^</sup> | 26.5 [26, 27]<br>(n=82) | 26 [25, 26]<sup>a</sup><br>(n=134) | 28 [28, 29]<sup>^,a,d</sup><br>(n=172) | 27 [26, 28]<sup>d</sup><br>(n=61) | 27 [26, 28]<br>(n=53) |
| **Sensory/Motor** |  |  |  |  |  |
| MDS-UPDRS III | 18 [16, 23]<br>(n=109) | 18.5 [16, 21]<br>(n=240) | 19 [18, 22]<br>(n=196) | 14 [12, 20]<br>(n=115) | 19 [13, 26] (n=60) |
| **Core Symptoms** |  |  |  |  |  |
| Prevalence of VH* | 29.6%<br>(n=71)<sup>^</sup> | 47.7%<br>(n=218)<sup>a</sup> | 17.7%<br>(n=158)<sup>a,b</sup> | 47.8%<br>(n=67)<sup>b</sup> | 26.3%<br>(n=38) |
| Prevalence of RBD (in PD<br>and DLB) | 80.6%<br>(n=36) | 77.3%<br>(n=172) | 57.6%<br>(n=85) | 70.5%<br>(n=44) | 76.5%<br>(n=34) |
| Prevalence of cognitive<br>fluctuations | 52.9%<br>(n=34) | 63.7%<br>(n=157) | 54.8%<br>(n=31) | 52.5%<br>(n=40) | 57.9%<br>(n=19) |
| **Cognitive Status (Baseline)** |  |  |  |  |  |
| PD-MCI (%; n=59) | 25.42%<br>(n=15) | 28.81%<br>(n=17) | 28.81%<br>(n=17) | 13.55%<br>(n=8) | 3.39%<br>(n=2) |
| PDD (%; n=33) | 9.09%<br>(n=3) | 27.27%<br>(n=9) | 21.21%<br>(n=7) | 39.39%<br>(n=13) | 3.03%<br>(n=1) |
| **Striatal Binding Ratios (PD)** |  |  |  |  |  |
| Total Striatum | 0.66 [0.52, 0.88]<br>(n=18) | 0.61 [0.55, 0.80]<br>(n=25) | 0.73 [0.62, 0.79]<br>(n=32) | 0.625 [0.490, 0.790]<br>(n=10) | 0.76 [0.68, 0.84]<br>(n=2) |
| Total Caudate | 0.66 [0.54, 0.93]<br>(n=18) | 0.72 [0.50, 1.03]<br>(n=25) | 0.71 [0.59, 0.84]<br>(n=32) | 0.70 [0.52, 0.92]<br>(n=10) | 0.71 [0.49, 0.93]<br>(n=2) |
| Total Putamen | 0.53 [0.43, 0.75]<br>(n=18) | 0.50 [0.45, 0.65]<br>(n=25) | 0.67 [0.57, 0.73]<br>(n=32) | 0.50 [0.39, 0.66]<br>(n=10) | 0.72 [0.68, 0.77]<br>(n=2) |
| **αSyn SAA positivity** | 93.8% (n=32) | 82.8% (n=29) | 84.8% (n=46) | 100% (n=20) | 50% (n=4) |
| **Stage\*<sup>^</sup> (Mean ± SD)** | 0 | 12.3 ± 11.18<sup>a</sup> | 8.01 ± 5.80<sup>a</sup> | 9.74 ± 7.33 | 8.42 ± 7.07 |

To enable robust subtype characterisation across a transdiagnostic cohort (PD, PD-MCI, PDD, DLB, and iRBD), subtype naming was anchored to severe atrophy events (z>2), as these represent the most biologically discriminative points of divergence between subtypes, whereas earlier low-amplitude changes were frequently shared across trajectories. The largest classified Subtype ‘A’ (Early cortico-limbic/late basal ganglia; n=297, 41.8%) presented early simultaneous neocortical atrophy along with amygdala and hippocampal degeneration before progressing to subcortical structures. Subtype ‘B’ (Early basal ganglia/late limbic; n=215, 30.3%) showed mild cortical atrophy initially, but severe degeneration was seen earliest in the basal ganglia, before extending to thalamus, brainstem, and limbic structures. Subtype ‘C’ (Early temporo-limbic/late basal ganglia; n=123, 17.3%) was defined by early atrophy of the temporal lobe, cingulate and other limbic regions, with delayed basal ganglia involvement. The smallest Subtype, ‘D’ (Early basal ganglia–cingulate/late cortex; n=75, 10.6%), exhibited early basal ganglia atrophy along with posterior cingulate, followed by sequential involvement of limbic and neocortical areas. Demographic and clinical characteristics for the LBD subtypes are in Table 1. Striatal dopaminergic function, α-synuclein SAA status, and AD biomarker characterisation across the identified subtypes in an early-stage PD subset from the PPMI cohort are reported in the Supplementary Results and Supplementary Table S4.

### Subtypes exhibit distinct cortical atrophy profiles across SuStaIn stages

We next examined whether the stage-dependent pattern of atrophy varied between subtypes. Total cortical volumes (extracted using the Desikan-Killiany parcellation) were normalised against healthy controls and used as the outcome in linear models with stage, subtype, stage × subtype interaction, age, and sex as predictors. Across stages, Subtype B (Early basal ganglia/late limbic) showed relatively preserved volumes (β (Stage)=–0.026, 95% CI [–0.037, –0.015], *p_FDR_*<0.001). Using Subtype B as the reference, stage × subtype interaction terms indicated more widespread cortical atrophy across SuStaIn stages in Subtype A (Early cortico-limbic/late basal ganglia, β=0.053, 95% CI [0.041, 0.065], *p_FDR_*<0.001), Subtype C (Early temporo-limbic/late basal ganglia, β=0.083, 95% CI [0.066, 0.099], *p_FDR_*<0.001), and Subtype D (Early basal ganglia–cingulate/late cortex, β=0.048, 95% CI [0.028, 0.068], *p_FDR_*<0.001).

### Clinical diagnoses map differentially onto subtypes

Clinical diagnoses were unevenly distributed across the four subtypes (Table 1 and Figure 2). Excluding the unclassified patients, two-thirds (66.1%) of individuals with DLB were classified into Subtype A (Early cortico-limbic/late basal ganglia), which also represented the most common clinical diagnostic group within this subtype (53.9% of Subtype A) (Figure 2B). Half of patients with PD (49%) were classified into Subtype B (Early basal ganglia/late limbic), with PD diagnosis comprising most of this subtype (82.3% of Subtype B). While less common overall, Subtype C (Early temporo-limbic/late basal ganglia) and Subtype D (Early basal ganglia–cingulate/late cortex) had higher representation of individuals with iRBD, and each comprised similar proportions of patients with PD and DLB (Figure 2).

**Figure 2:**
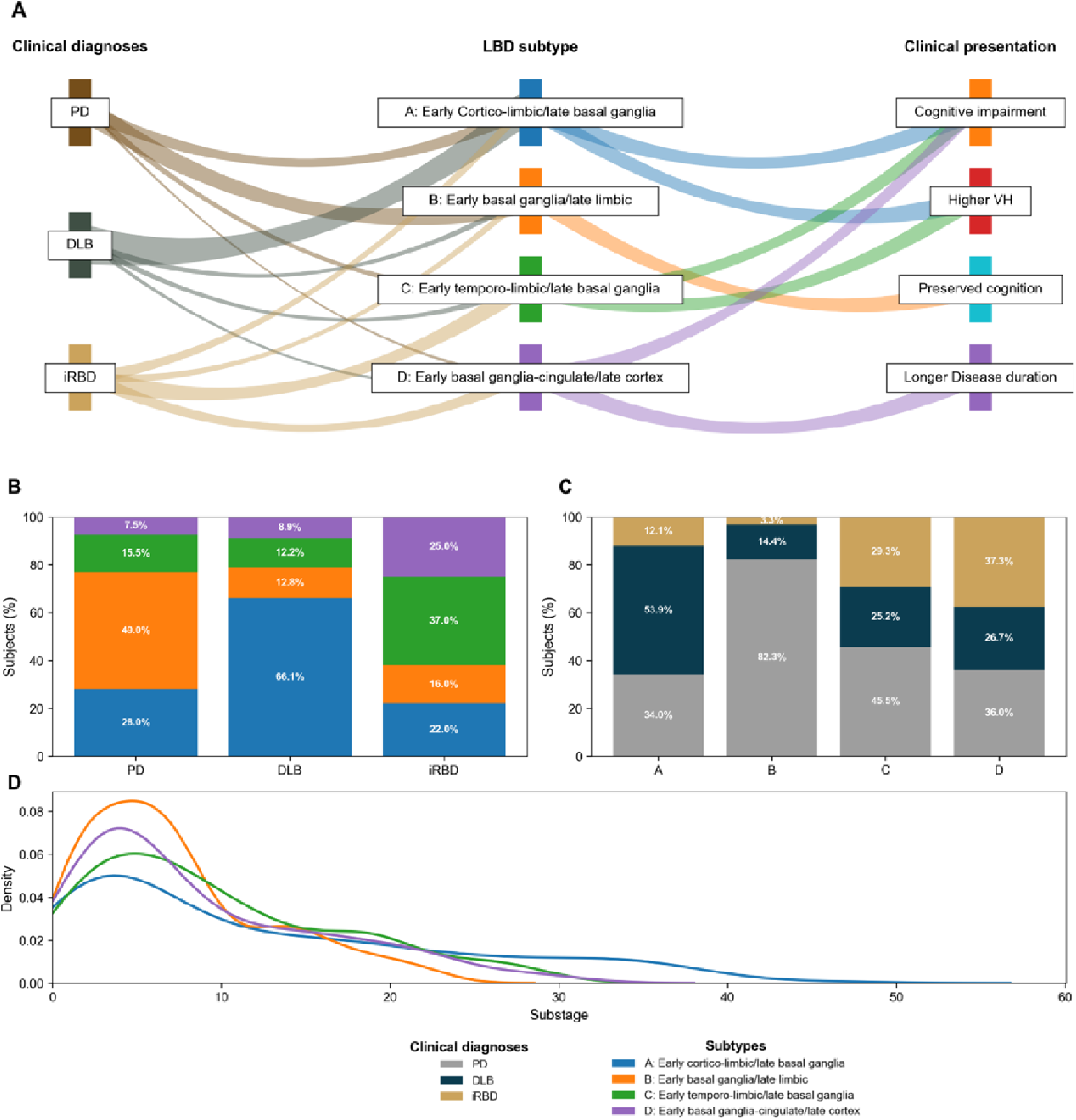
LBD subtypes cut across clinical diagnoses and align with clinical presentations. Subtype membership varies by diagnosis and explains symptom profiles beyond diagnosis alone. **A:** Profiles linking diagnosis to subtype and clinical presentations. Line widths from left to centre reflect proportions. Subtypes A and C link to higher visual hallucinations and lower cognition. Subtype B links to preserved cognition. Subtype D has higher disease duration. **B:** Subtype composition within each diagnosis (stacked bars, percentages). **C:** Diagnosis composition within each subtype (stacked bars, percentages). **D:** Probability density of number of patients in each SuStaIn stage by subtypes (kernel density plot). Subtypes: Blue = Subtype A (Early cortico-limbic/late basal ganglia), Orange = Subtype B (Early basal ganglia/late limbic), Green = Subtype C (Early temporo-limbic/late basal ganglia), Purple = Subtype D (Early basal ganglia–cingulate/late cortex). Clinical Diagnoses: Grey = Parkinson’s Disease, Teal = Dementia with Lewy Bodies, Yellow = Idiopathic REM Sleep Behaviour Disorder. PD, Parkinson’s disease. DLB, dementia with Lewy bodies. iRBD, idiopathic REM sleep behaviour disorder. VH, visual hallucinations.

### Relationship between SuStaIn stage and disease duration

The relationship between SuStaIn stage and disease duration since diagnosis differed across the subtypes. Subtype B had significantly lower mean stage (8.01±5.80) than Subtype A (12.30±11.18; p*_FDR_*<0.001) and C (9.74±7.33, p*_FDR_*<0.001). We examined whether SuStaIn stage, reflecting cumulative brain atrophy, was associated with disease duration by fitting a regression model with SuStaIn stage as the outcome, disease duration (years) as the main predictor, and subtype allocation and subtype probability as covariates. Independent of subtype, SuStaIn stage increased with disease duration in PD (β=0.167, 95% CI [0.024, 0.310], p*_FDR_*<0.042), with a trend in iRBD (β=0.455, 95% CI [−0.199, 1.110], p*_FDR_*=0.115), and the relationship was not significant in DLB (β=−0.039, 95% CI [-0.071, 0.007], p*_FDR_*=0.341). Subtype-specific, transdiagnostic analyses showed significant positive correlations between stage and disease duration, independent of clinical diagnosis, in Subtype B (Early basal ganglia/late limbic, β = 0.264, 95% CI [0.057, 0.471], *p_FDR_*=0.013), and C (Early temporo-limbic/late basal ganglia, β=0.514, 95% CI [0.023, 1.005], *p_FDR_*=0.040), but not in D (Early basal ganglia–cingulate/late cortex, β=0.112, 95% CI [–0.441, 0.664], *p_FDR_*=0.687). Interestingly, in Subtype A (Early cortico-limbic/late basal ganglia), stage was inversely correlated with disease duration (β=–0.328, 95% CI [–0.647, –0.009], *p_FDR_*=0.044), which can be suggestive of either more aggressive atrophy accumulation and/or delayed diagnosis.

### Subtypes capture cognitive and motor heterogeneity

We next compared cognitive and motor performance across subtypes using MMSE (n=427) and MDS-UPDRS-III scores (n=611), respectively (Figure 3A and 3B). At the group level, Subtype B (Early basal ganglia/late limbic) showed higher MMSE than Subtypes A (Early cortico-limbic/late basal ganglia, p*_FDR_*<0.001), C (Early temporo-limbic/late basal ganglia, p*_FDR_*=0.004), and D (Early basal ganglia–cingulate/late cortex, p*_FDR_*=0.006), consistent with its milder atrophy profile. Motor severity measured using MDS-UPDRS-III did not differ across subtypes when controlling for SuStaIn stage.

**Figure 3:**
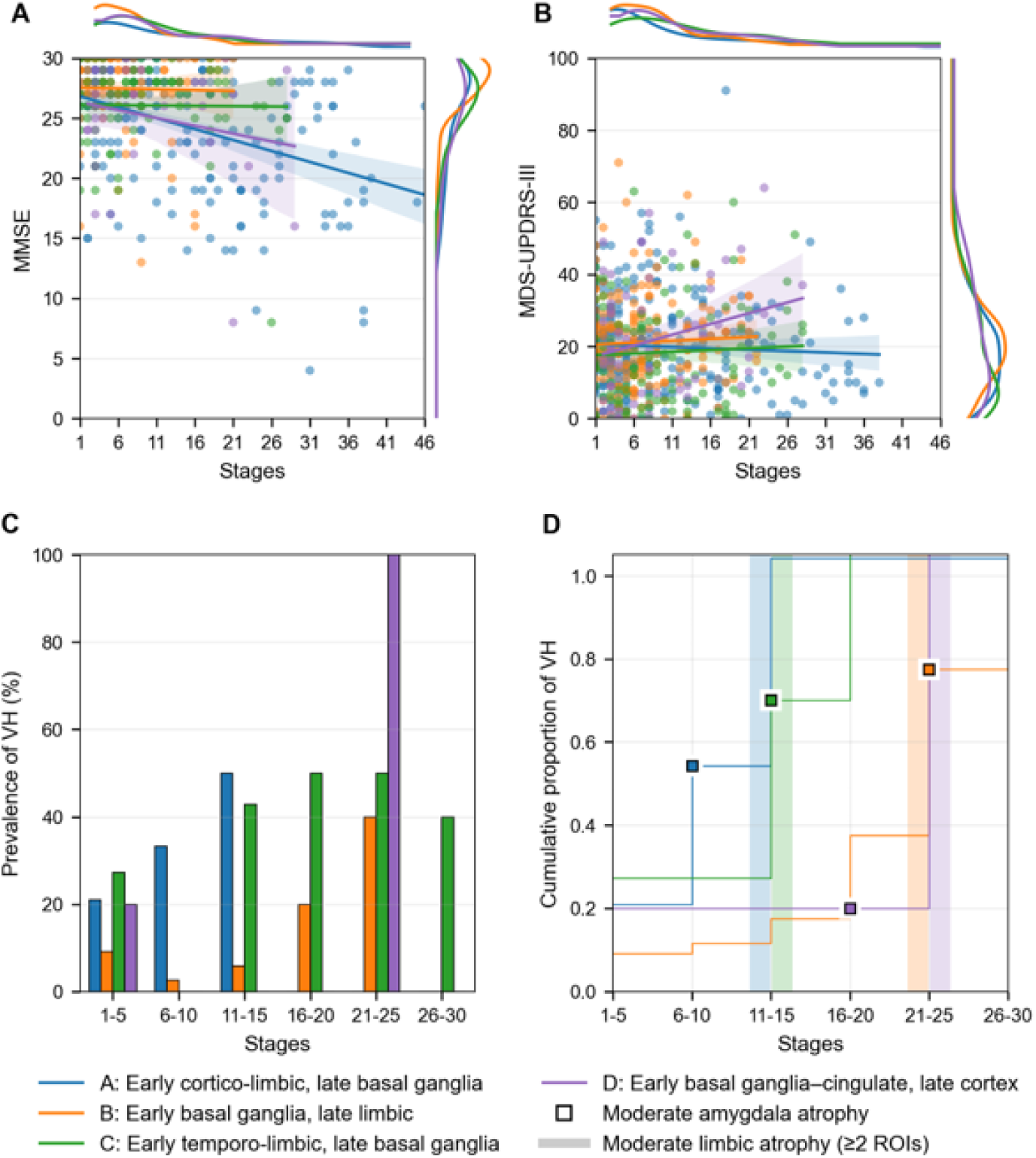
Subtype trajectories of clinical features in Lewy body diseases. SuStaIn-derived subtypes show distinct trajectories of cognition and motor decline, and differential timing of visual hallucinations. **A–B:** Cognitive (A) and motor (B) trajectories across stages for all four transdiagnostic subtypes, measured by MMSE and MDS-UPDRS-III, respectively (linear regression, adjusted for age and sex). Shaded regions denote 95% confidence intervals; marginal density plots display data distributions. **C:** Prevalence of visual hallucinations across SuStaIn stage bins in PD, stratified by SuStaIn derived subtypes. **D:** Cumulative emergence of visual hallucinations within PD subtypes. Lines depict the proportion of individuals in each subtype who have experienced hallucinations by the end of each stage bin. Square markers indicate moderate amygdala atrophy (z > 1). Vertical shaded bands indicate the subtype-specific stage range at which the limbic involvement threshold is reached, defined as moderate atrophy, z > 1, in at least two of the four modelled limbic ROIs. Blue = Subtype A (Early cortico-limbic/late basal ganglia), Orange = Subtype B (Early basal ganglia/late limbic), Green = Subtype C (Early temporo-limbic/late basal ganglia), Purple = Subtype D (Early basal ganglia–cingulate/late cortex). PD, Parkinson’s disease, MMSE Mini-Mental State Examination, MDS-UPDRS III, Movement Disorders Society Unified Parkinson’s Disease Rating Scale Section III.

Across the cohort, MMSE declined with advancing stage (β=–0.16, 95% CI [−0.22, −0.09], p*_FDR_*<0.001), consistent with worsening cognitive performance over disease progression. However, subtype × stage interaction was not significant for cognition, which suggests that the spatiotemporal signature of atrophy (i.e., subtype) may be less important than overall severity of atrophy when prognosing general cognitive decline. By contrast, motor severity increased more steeply with advancing stage in Subtype C (β=0.54, 95% CI [0.05, 1.03], *p_FDR_*=0.030) and showed a trend in Subtype D (β=0.68, 95% CI [–0.06, 1.43], *p_FDR_*=0.070), both relative to Subtype A.

We investigated autonomic symptom burden in PD including pain, urinary dysfunction, constipation, light-headedness, and fatigue, using pairwise logistic regression adjusted for age, sex, and SuStaIn stage. Symptoms were dichotomised as clinically significant at scores greater than 2. Patients with PD in Subtype B showed reduced odds of fatigue compared with Subtype A (OR=0.18, 95% CI [0.03, 0.94]; p*_FDR_*=0.041), indicating that the patients with PD in the malignant Subtype A reported higher fatigue.

### Hallucinations associate with limbic trajectories and amygdala atrophy

Across the classified transdiagnostic LBD subtypes (irrespective of clinical diagnosis), the prevalence of visual hallucinations was highest in Subtype A (Early cortico-limbic/late basal ganglia, 47.7%) and C (Early temporo-limbic/late basal ganglia, 47.8%) and lowest in early basal ganglia subtypes B (Early basal ganglia/late limbic, 17.7%) and D (Early basal ganglia–cingulate/late cortex, 26.3%). Accounting for stage, subtypes with early limbic degeneration (A and C together) showed significantly higher prevalence of hallucinations than the early basal ganglia subtypes (B and D) (OR=3.54, 95% CI [2.29, 5.50], Wald χ^2^=31.98, p*_FDR_*<0.001).

Transdiagnostic subtypes revealed heterogeneity of visual hallucinations even within a particular clinical diagnostic group. Focusing on PD, where hallucinations are not a core feature of diagnosis and may develop at any time in the disease, we found that participants with PD classified to Subtypes A (p*_FDR_*<0.001; Wald’s χ^2^=5.24) and C (p*_FDR_*=0.001; Wald’s χ^2^=4.41) showed significantly higher prevalence than Subtype B (Figure 3C). Participants with PD in these subtypes also demonstrated an earlier rise in terms of SuStaIn-stage of visual hallucinations compared to subtypes B and D. Patients were further stratified into pre-and post-limbic phases based on the subtype-specific stage at which limbic regions became involved (Subtype A: stage 10; Subtype B: stage 22; Subtype C: stage 10; Subtype D: stage 18). Visual hallucinations were significantly more frequent (Figure 3D) following this transition after adjusting for age, sex and subtype (Wald χ²=5.34, p=0.021). To assess whether this association was specific to limbic involvement, we performed additional analyses examining the temporal relationship between hallucination prevalence and atrophy in other cortical and subcortical regions, which did not demonstrate a consistent stage-dependent relationship (Supplementary Figure S4). Qualitatively, this rise coincided in particular with moderate atrophy in the amygdala (defined by a z-score of 1), highlighting it as a potentially important region of interest (Figure 3D).

Individuals with PD experiencing visual hallucinations had significantly lower amygdala volume (p*_FDR_*<0.001), and this was confirmed with a post-hoc analysis assessing cumulative prevalence of VH, which demonstrated a steep increase in visual hallucinations once amygdala atrophy exceeded a z-score threshold of 1 in subtypes A, C and D. To test the added benefit of data-driven subtypes to amygdala volume across Parkinson’s disease, we fitted three stage-adjusted logistic models predicting visual hallucinations. Model 1 included amygdala z-score as a fixed effect (β=−0.43, SE=0.19, OR=0.65, 95% CI [0.45, 0.95], p*_FDR_*=0.026) achieving an AUC of 0.62 (95% CI [0.52, 0.70]). Model 2 included subtype as a sole predictor (AUC=0.66, 95% CI [0.57, 0.75]). Using Subtype B as reference, Subtype C showed the strongest association with VH (OR=5.09, 95% CI [1.89, 13.70], p*_FDR_*=0.004), while Subtype A showed an elevated risk (OR=2.24, 95% CI [0.98, 5.10], p*_FDR_*=0.045). Model 3 included amygdala volume, subtype and SuStaIn stage and showed improved discrimination (AUC=0.73, 95% CI [0.64, 0.81]). In this model, subtype remained independently associated with VH after adjustment for amygdala volume, with Subtype C (β=1.18, SE=0.55, OR=3.25, 95% CI [1.10, 9.61], p*_FDR_*=0.033) and Subtype A (β=0.92, SE=0.46, OR=2.51, 95% CI [1.03, 6.13], p*_FDR_*=0.044) showing elevated VH risk relative to Subtype B. Reduced amygdala volume remained an independent predictor (β=−0.47, SE=0.21, OR=0.63, 95% CI [0.42, 0.95], p*_FDR_*=0.027), and more advanced disease stage was independently associated with increased VH risk (β=0.07, SE=0.03, OR=1.07, 95% CI [1.01, 1.13], p*_FDR_*=0.033).

All three models significantly improved over a null intercept-only model, with the combined Model 3 achieving the highest discriminatory performance (Model 1: χ²=5.26, p*_FDR_*=0.022, AUC=0.62, 95% CI [0.52, 0.70]; Model 2: χ²=12.29, p*_FDR_*=0.007, AUC=0.66, 95% CI [0.57, 0.75]; Model 3: χ²=21.52, p*_FDR_*<0.001, AUC=0.73, 95% CI [0.64, 0.81]). Model 3 significantly outperformed Model 1 (χ²=16.26, p=0.003), confirming that the joint consideration of both spatial pattern and temporal progression provides predictive information for VH risk beyond amygdala atrophy alone.

### Data-driven subtypes predict the development of cognitive impairment in PD

Participants with PD were distributed across all subtypes (Figure 2). We next examined whether global cognition in PD varies by subtype and whether certain subtypes carry a greater risk of developing cognitive impairment (MoCA < 24). For this analysis, we used the available subsample (30%) of PD participants with longitudinal measures (n=135; median follow-up=4.9 years [3.1, 5.5]; age=65.1, 95% CI [64, 67.2]; baseline MoCA=27.00, 95% CI [27, 28]). Detailed demographics are provided in Supplementary Table S5.

Analyses included individuals with a confirmed subtype and ≥ 1 follow-up MoCA score (excluding stage 0, and subtype probability < 0.50). Log-rank tests indicated a significant overall difference in survival curves across subtypes (χ²=8.74, p=0.033). Pairwise comparisons indicated that Subtype A had a significantly faster rate of conversion to MCI than Subtype B (χ²(1)=8.61, p*_FDR_*=0.020), while no other pairwise comparisons reached significance after FDR correction. The Cox proportional hazards model adjusted for age, sex and stage also showed a statistically significant fit (Log likelihood χ² (6)=14.99, p=0.021), and the proportional hazards assumption was confirmed using Schoenfeld residuals (p>0.05). Stage was not significantly associated with time to conversion to MCI (Hazard ratio (HR)=0.99, 95% CI [0.94, 1.05], p=0.64). Compared to Subtype B, Subtype A exhibited a significantly higher risk of conversion (HR=2.91, 95% CI[[[1.52, 5.59], p*_FDR_*=0.007), while Subtypes C and D showed elevated but non-significant risks (Subtype C: HR=1.77, 95% CI [0.80, 3.92], p*_FDR_*=0.237; Subtype D: HR=1.79, 95% CI [0.68, 4.67], p*_FDR_*=0.279). The cumulative number of events for each subtype over time, and the number of patients at risk (not yet converted to MCI at that time point) are included in Figure 4. A logistic regression model including stage × subtype interaction, age, and sex as fixed effects predicted MCI with an AUC of 0.75, 95% CI [0.69, 0.81].

**Figure 4:**
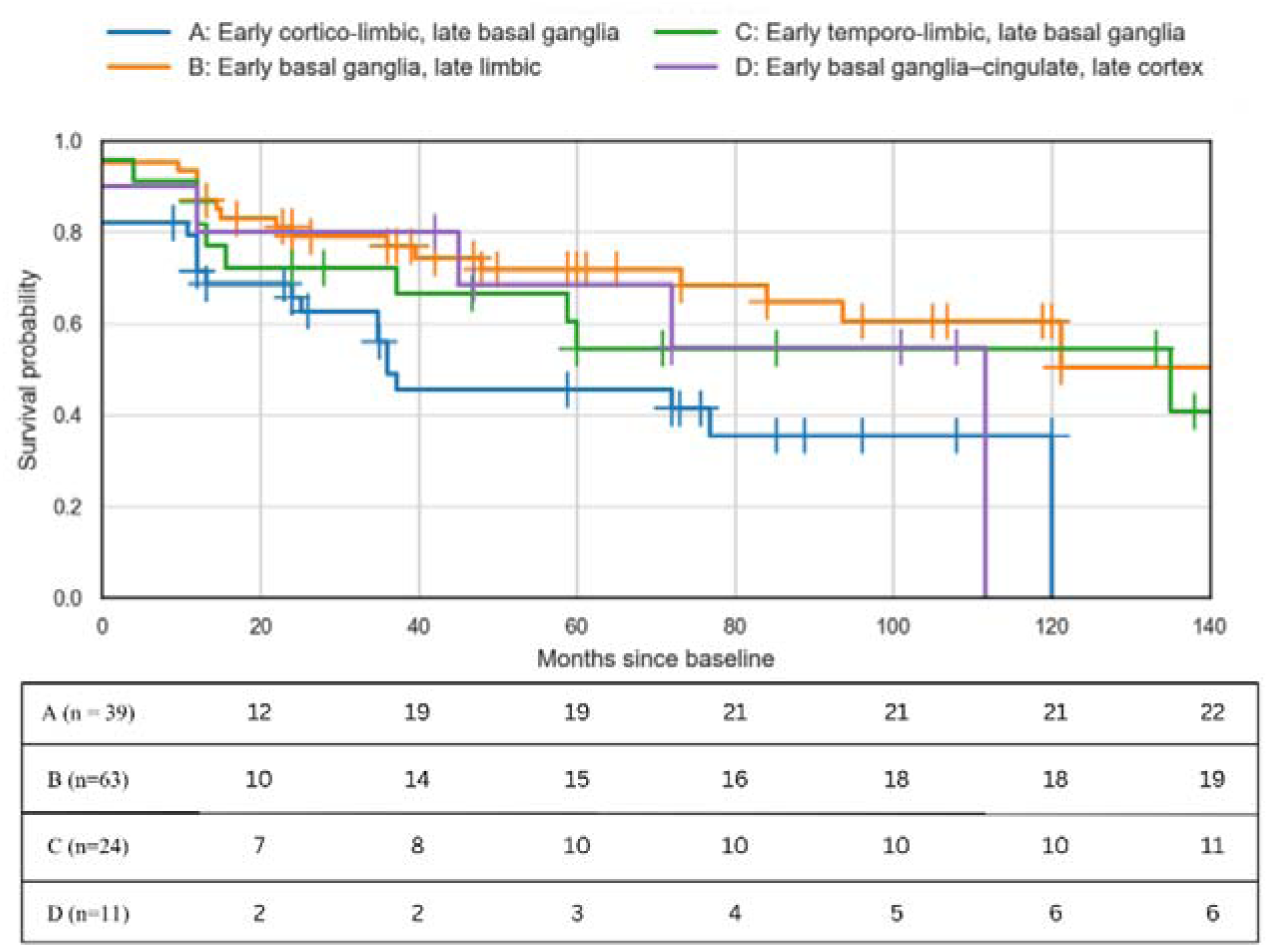
Cognitive outcomes in PD by subtype. Kaplan–Meier curves for time to MCI conversion by subtype using months since baseline (hazard ratios estimated using Cox proportional hazards regression, adjusted for age, sex and stage). Event (MCI) is MoCA < 24. Patients are censored at their last follow-up if they do not reach an event indicated by ‘+’ on the coloured lines. Blue = Subtype A (Early cortico-limbic/late basal ganglia), Orange = Subtype B (Early basal ganglia/late limbic), Green = Subtype C (Early temporo-limbic/late basal ganglia), Purple = Subtype D (Early basal ganglia–cingulate/late cortex). MoCA, Montreal Cognitive Assessment. MCI, mild cognitive impairment. PD, Parkinson’s disease.

### Subtypes exhibit differential clinical presentations over time

In longitudinal RBDSQ analyses in individuals with PD experiencing RBD symptoms (RBDSQ ≥ 5 at baseline), there were no main-effect differences between subtypes. Detailed demographics are provided in Supplementary Table S5. Interaction terms with time indicated faster RBDSQ worsening for Subtype C (Early temporo-limbic/late basal ganglia) compared with Subtype A (Early cortico-limbic/late basal ganglia) (β=−0.03, 95% CI [−0.05, −0.01], p*_FDR_*=0.010) and Subtype B (Early basal ganglia/late limbic) (β=−0.04, 95% CI [−0.06, −0.01], p*_FDR_*<0.001) (Figure 5). No difference in MDS-UPDRS-III or MoCA scores was observed.

**Figure 5:**
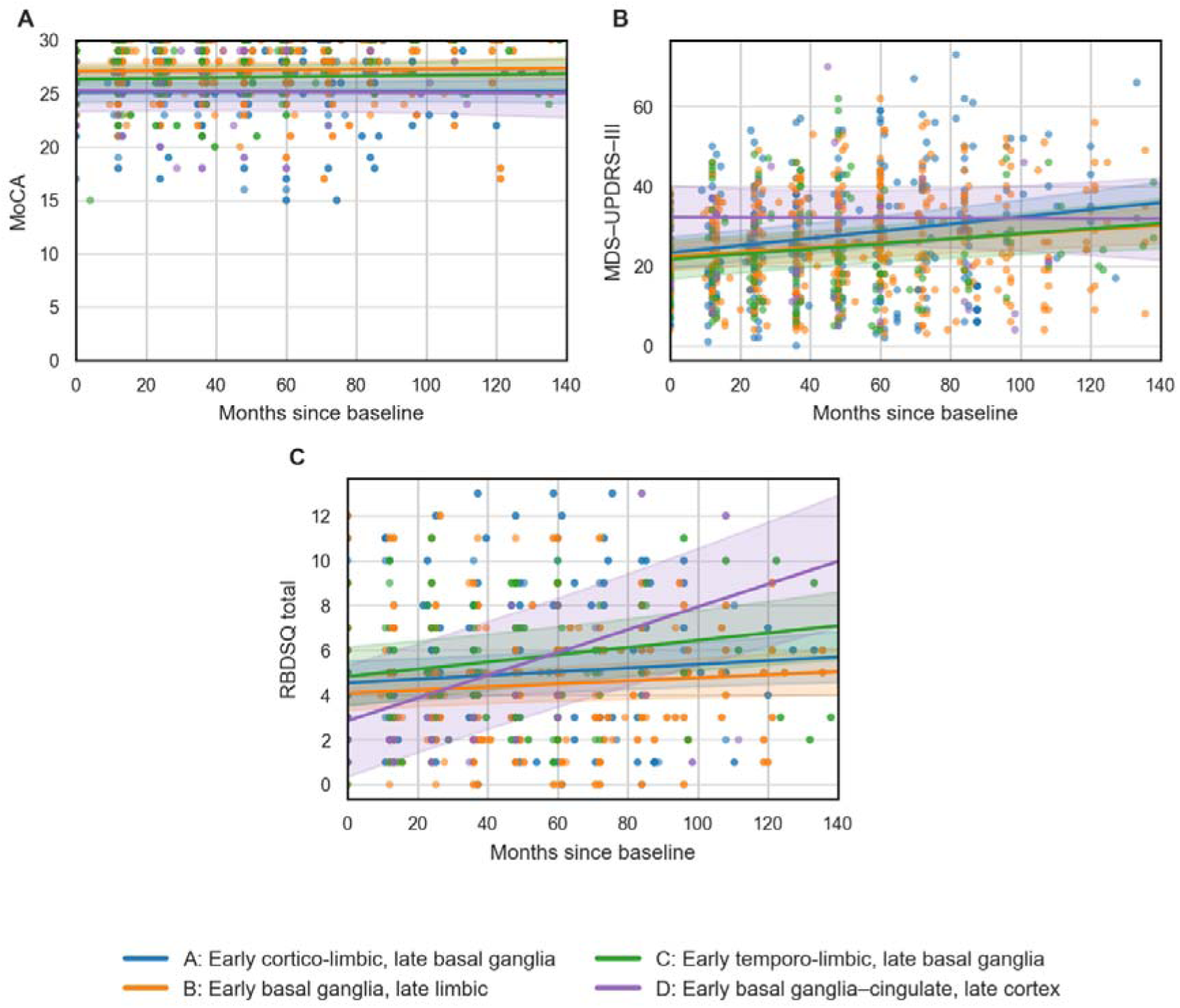
Longitudinal motor severity and RBD symptoms by subtype. Linear mixed-effects models show subtype-specific slopes over time. **A:** MoCA versus months since baseline by subtype. Lines are mixed effects estimates with 95% confidence intervals, adjusted for age and sex. MDS-UPDRS III versus months since baseline by subtype with model estimates and 95% confidence ribbons. **B:** RBDSQ total versus months since baseline by subtype with model estimates and 95% confidence ribbons. Individuals having RBDSQ ≥ 5 are only included. Blue = Subtype A (Early cortico-limbic/late basal ganglia), Orange = Subtype B (Early basal ganglia/late limbic), Green = Subtype C (Early temporo-limbic/late basal ganglia), Purple = Subtype D (Early basal ganglia–cingulate/late cortex). MDS-UPDRS III, Movement Disorder Society Unified Parkinson’s Disease Rating Scale part III; RBDSQ, REM Sleep Behaviour Disorder Screening Questionnaire.

### SuStaIn stage predicts dementia conversion in iRBD

Prevalence of RBD varied across the transdiagnostic LBD subtypes. Subtype B (Early basal ganglia/late limbic, 57.6%) showed the lowest RBD prevalence and, after adjusting for stage, showed significantly lower prevalence than A (Early cortico-limbic/late basal ganglia, 77.3%; Subtype A vs B: p*_FDR_*<0.001; Wald’s χ^2^=10.6), C (Early temporo-limbic/late basal ganglia, 70.5%; Subtype B vs C: p*_FDR_*<0.001; Wald’s χ^2^=11.1) and D (Early basal ganglia–cingulate/late cortex, 76.5%; Subtype B vs D: p*_FDR_*<0.001; Wald’s χ^2^=11.1). Stage-related prevalence of RBD is mentioned in Supplementary Results and Supplementary Figure S3.

Longitudinal phenoconversion data were available for the iRBD cohort from Sydney (n=41, median follow-up time of 3.58 years, 95% CI [1.58, 4.41]; detailed demographics in Supplementary Table S6). During follow-up, 10 patients with iRBD converted to PD and 7 to DLB, while 24 remained unconverted. Among converters, 6/10 iRBD-PD converters (60%) were classified into Subtype D (Early basal-ganglia–cingulate/late cortex, binomial p*_FDR_*=0.020), whereas 4/7 iRBD-DLB converters (57.1%) were classified into Subtype A (binomial p*_FDR_*=0.027) (Supplementary Figure S5).

To test whether disease severity predicted time to phenoconversion in iRBD, we fitted Cox proportional hazards models with time to conversion as the outcome and SuStaIn stage, age, and subtype as covariates. Models showed no significant association between stage and conversion time across the full LBD cohort or among PD converters. In contrast, a higher stage was associated with quicker phenoconversion to DLB (HR=1.45, 95% CI [1.03, 2.03], *p_uncorrected_*=0.030, Cliff’s δ=0.50) but did not survive *FDR* correction. In all three models, subtype was not associated with time to phenoconversion, which may be due to the limited number of phenoconverters within each subtype.

### Individual trajectories of atrophy within PD and DLB

Finally, to compare with prior diagnosis-specific studies in PD and to validate the combined LBD findings, we modelled PD and DLB separately. In DLB (n=275), three trajectories emerged (Supplementary Figure S6): DLB-A (50%), with early neocortical, and cingulate atrophy extending to limbic and subcortical structures (consistent with a rostro-caudal pattern); DLB-B subtype (34%) with early neocortical and limbic involvement (entorhinal cortex, amygdala, and hippocampus) followed by the cingulate and thalamus; and DLB-C (16%) with early caudate atrophy followed by progressive involvement of amygdala, hippocampus, accumbens and cortex.

Three distinct patterns also emerged in PD (n=436): PD-A (36%), with early diffuse cortical atrophy followed by the limbic and basal ganglia; PD-B (35%) subtype, with early atrophy in the amygdala, accumbens, cingulate; and PD-C (29%) with early degeneration of the entorhinal cortex, followed by prominent basal ganglia, thalamus, and brainstem involvement prior to broader cortical atrophy (Supplementary Figure S7). Clinical differences across these diagnosis-specific models are described in Supplementary Results, Supplementary Tables S7-8, and Supplementary Figure S8.

## Discussion

In this multicentre imaging study, we applied an unsupervised machine learning-based disease-progression model and identified four transdiagnostic subtypes of atrophy based on brain volume features extracted from T1-weighted MRI scans in a large cohort of individuals with clinical PD, DLB, and iRBD. These spatiotemporal patterns of atrophy aligned with distinct but overlapping clinical phenotypes and demonstrated prognostic value for phenoconversion in iRBD and the risk of progression to cognitive impairment in PD. Separately, we applied the same method within diagnostic groups, which recapitulated PD neurodegenerative subtypes described previously and highlighted the DLB-specific atrophy trajectories.

To date, the majority of prior research has examined LBD cross-sectionally as discrete clinical syndromes.^42,43^ A key advance of our work lies in extending this unbiased biological framework across a large sample of PD and DLB alongside individuals with prodromal iRBD, reflecting a wider variety of LBD cases at different clinical and pathological stages. Our combined model identified four transdiagnostic trajectories with distinct clinical phenotypes demonstrating broad alignment with known clinical/diagnostic subtypes, thereby capturing additional spatiotemporal granularity in disease expression, and supporting biological stratification. Subtype A (Early cortico-limbic/late basal ganglia) was dominated by individuals with DLB and was associated with lower baseline global cognition and higher risk of developing MCI in PD.^52,53^ In contrast, Subtype B (Early basal ganglia/late limbic) concentrated participants with PD, with preserved cognition and lower cortical atrophy at baseline, lower prevalence of visual hallucinations and RBD, and, on average, more stable cognition at follow-up. As expected, the loss of dopamine axons in the basal ganglia was not reflected in significant atrophy in this or any subtype,^54–56^ but has been shown to correlate with a preclinical reduction of neuromelanin rather than volume.^57^ Subtypes A and B complement previous data-driven and hypothesis-driven subtyping efforts in PD, which have usually emphasised at least two subtypes: one with a more aggressive course with earlier cognitive impairment such as the ‘diffuse malignant’ subtype (most similar to Subtype A); and a slower-progressing mild motor-predominant subtype^7^ (most similar to Subtype B). In addition, our model also revealed two further trajectories in Subtypes C (Early temporo-limbic/late basal ganglia) and D (Early basal ganglia–cingulate/late cortex), which concentrated patients with iRBD. These subtypes may be interpreted as more granular spatiotemporal variants within broader cortico-limbic and basal ganglia-weighted patterns of degeneration, respectively. Of interest, Subtype C which had relatively early brainstem and thalamic atrophy, showed faster progression of RBDSQ symptoms over time.

Two clinical iRBD subtypes have also been previously identified, broadly characterised as cortical-first and subcortical-first trajectories, with the former more frequently associated with subsequent DLB phenoconversion.^9,58^ Interestingly, in our study, Subtypes C and D were enriched for individuals with iRBD but also included participants with manifest PD and DLB, suggesting that they may capture additional spatiotemporal variation across the LBD spectrum rather than representing strictly prodromal atrophy patterns. Subtype D, for example, was associated with longer disease duration, early basal ganglia degeneration, and relatively steeper motor decline. In line with this, a slowly progressive DLB neurodegenerative subtype with prodromal iRBD has been recently identified.^59^ Similar intermediate subtypes have also been derived from other cross-sectional clinical and pathological cohorts.^7,12,60^ Altogether, our results demonstrate that imaging-derived subtypes capture clinically meaningful heterogeneity across diagnoses.

In addition to subtyping, data-driven staging added complementary insights into disease course. Subtype A (strong dementia subtype) showed an inverse association between spatial extent of atrophy and disease duration, possibly driven by individuals with DLB having simultaneously advanced atrophy and diagnosis indexed by dementia (which is a potentially late event in the disease course, as opposed to MCI or iRBD), whereas Subtypes B and C showed the opposite: atrophy extent was positively associated with longer disease duration. The subtype-specific discrepancy between disease duration and biological severity underscores the limitations of ‘disease duration’ as a surrogate for disease stage transdiagnostically. Duration is typically measured from the time of diagnosis and is therefore influenced by recall bias, while overlooking potentially prolonged clinical or subclinical prodromal intervals. This is particularly relevant in DLB, which often manifests later than PD and may be preceded by a longer subclinical phase. Indeed, iRBD more frequently precedes DLB than PD, and may arise decades before the onset of dementia.^6^ Prior work comparing PD with and without RBD has reported limited differences, but longitudinal studies have shown greater frontotemporal atrophy in RBD-positive individuals, and consistent with this Subtypes A and C showed higher RBD burden and greater cortical involvement, while the low RBD prevalence in Subtype B (PD-dominated subtype) suggests that RBD is not uniformly a prodromal symptom across PD presentations. With efforts currently underway to develop unified clinical staging models for LBD, our findings highlight that the underlying neurodegeneration is heterogeneous and subtype-dependent. A combined clinical-atrophy staging model would offer a more biologically grounded basis for individualising disease progression across LBD.^6^

The imaging-derived subtypes identified in this study provide complementary insights and show partial correspondence with different SuStaIn subtypes derived in previous cross-sectional pathological studies.^13,14,61^ However, these pathological studies differ from the present study in their case selection, with the largest study published using all cases with Lewy pathology (n=814), of whom 66% having clinicopathological AD diagnosis, a further 11% had Lewy pathology with no clinical phenotype, and fewer than 25% had clinical LBD (n=187, the majority with PD aged 82 years on average).^14^ Despite this, the early amygdala and entorhinal pathological subtype ‘S1’, which concentrated their AD cases, parallels the limbic atrophy of our dementia-dominant subtype A, suggesting that this subtype is relevant for clinical LBD. The olfactory-brainstem pathological subtype S2 shares features with our iRBD-dominant Subtype C and there is some similarity between their brainstem-early/olfactory later (S3) and our Subtype D. Subtype B, which for us concentrated our largest cohort of individuals with PD (PD n=436), does not appear to have a direct pathological analogue, potentially due to the dispersal of their clinical PD cases between the three pathological subtypes observed. The second pathological study assessed only PD (n=54) plus preclinical LBD (n=2)^61^ and found two subtypes, a major subtype driven by brainstem then amygdala, limbic and finally neocortical Lewy pathology (consistent with Braak staging), and a minor subtype that was amygdala-driven (consistent with the dementia-dominant subtype A). The third study assessed 302 cases with Lewy pathology concentrating on incidental Lewy pathology cases to find the earliest origin sites for Lewy pathology,^13^ and so cannot be compared with our data of clinical LBD. The relative overlap between some pathology and imaging-derived subtypes suggests that early disease processes can be tracked forward to their end-stage manifestations, supporting the utility of *in vivo* atrophy patterns as biomarkers of progression. This convergence strengthens the case for imaging-based subtyping as a clinically meaningful complement to pathological staging, offering a framework that captures selective vulnerability and links it to measurable outcomes over time.

From a prognostic perspective, our model also provided insights into the development of cognitive impairment by identifying trajectories associated with increased risk of dementia across LBD. Subtype A was enriched for individuals with DLB and also showed the highest risk of MCI in PD, with over half of individuals with PD developing cognitive decline. This dementia-prone trajectory was characterised by extensive cortico-limbic atrophy, paralleling patterns reported in AD.^52,53^ These findings suggest that cortico-limbic regions constitute a convergence zone of vulnerability which can manifest with single or multiple proteinopathies. In line with this, amyloid-β and tau co-pathologies are frequent in both PDD and DLB,^62,63^ and their presence increases the risk of dementia in PD and modifies the interval between cognitive and motor symptoms in PD.^62,64^ Degeneration of cholinergic hubs has been associated with cognitive impairment in both PD^65^ and iRBD,^66^ supporting a broader role for cholinergic denervation across LBD.^31,67^ Such cholinergic atrophy is closely coupled to medial temporal grey matter loss, with hippocampal atrophy mediating the relationship between cholinergic atrophy and cognitive deficits.^68^ The spatiotemporal sequence of limbic degeneration could explain the pattern of cognitive symptoms and identify subtypes with greater cholinergic vulnerability. Building on prior work showing that cognitive impairment is common and can emerge early in PD,^69^ our results suggest that a subset of patients with PD share an atrophy trajectory with DLB and that SuStaIn subtyping can identify individuals at increased risk of cognitive decline.

Another clinically relevant finding of our model was that it stratified the direction of iRBD phenoconversion, providing a biologically grounded framework for predicting disease trajectory. Predicting phenoconversion of iRBD has been a major challenge due to marked heterogeneity and the absence of reliable biomarkers.^4,70,71^ Addressing that, our transdiagnostic model demonstrated predictive utility by differentiating individuals with iRBD according to their eventual manifest LBD diagnosis. Individuals with iRBD assigned to the limbic-dominant Subtype A were more likely to convert to DLB than to PD, and higher SuStaIn atrophy stages within this subtype were associated with faster phenoconversion. While these findings are based on a small, single-centre cohort and should be interpreted cautiously, they are broadly consistent with prior observations suggesting a higher likelihood of DLB conversion following cortical-dominant trajectories.^45^ By anchoring phenoconversion prediction to anatomically derived subtypes trained jointly with manifest PD and DLB, our approach integrates spatial and temporal disease information, explaining clinical variability and progression.

Beyond progression of neurodegeneration, the model also yielded pathophysiological insights into the core symptom manifestation of visual hallucinations. In this study, hallucinations were most frequent in limbic-dominant subtypes and emerged earlier along the inferred trajectory coinciding with development of moderate amygdala atrophy. Previous neuroimaging findings implicate the amygdala in the pathophysiology of visual hallucinations, reporting either bilateral reduction of amygdala volumes,^33^ or decreased functional connectivity between the amygdala and visual and attentional networks,^32^ suggesting the role of both structural and network-level contributions as well as heterogeneity across cohorts. Pathological evidence also implicates amygdala Lewy pathology in the manifestation of visual hallucinations in PD.^72–75^ While visual hallucinations likely reflect distributed network dysfunction involving visual, attentional, and subcortical systems, we focused on the amygdala as a biologically grounded and reproducible inflection point within this broader process. Subtyping using all clinical LBD captured this variability more clearly, with Subtype B showing a lower prevalence of hallucinations and preserved amygdala volumes compared to other subtypes. Notably, when accounting for subtype-specific progression, visual hallucinations were significantly more frequent following this transition, identifying limbic involvement as a critical stage-dependent inflection point for hallucination risk. Across subtypes, the proportion of participants with VH rose dramatically once stages reflecting limbic involvement were reached, and once amygdala atrophy reached a specific threshold. The convergence of these results across imaging modalities and neuropathological studies supports existing mechanistic frameworks^34,75^ in which amygdala degeneration increases susceptibility to visual hallucinations but also accounts for the variability in its effect. Importantly, our model demonstrates that combining amygdala atrophy with SuStaIn-derived subtype and stage improves discrimination of hallucination risk compared to amygdala volume alone. Clinically, this indicates that hallucination risk is not determined by regional atrophy alone, but by its position within the disease trajectory, enabling more precise identification of patients at higher risk and informing monitoring strategies and earlier targeted interventions.

In addition to the transdiagnostic modelling, we also recapitulated subtyping within clinical syndromes. In our analyses, the PD-only model reproduced the cortical (DLB-like), limbic and subcortical trajectories described previously,^42,43^ whereas the DLB-only model revealed trajectories with greater cortical involvement. Early hippocampal atrophy was observed only in the DLB-A subtype, while pronounced striatal atrophy was confined to DLB-C. However, these subtypes were not accompanied by group-level differences in cognition within either the PD or DLB syndromes, nor by differences in motor impairment in DLB. Across diagnoses, early caudate atrophy was evident in DLB, while PD showed combined caudate-putamen degeneration; this was reflected in DLB-C and PD-C, which exhibited earlier striatal involvement than other subtypes. These findings help reconcile certain inconsistencies in the literature, such as variable reports of hippocampal involvement^76^ in prodromal DLB and heterogeneity in striatal involvement.^77^ Consistent with prior work, diagnosis-specific models provided limited insights as overlapping phenotypes were not considered, underscoring the need for transdiagnostic approaches to resolve such phenotypic heterogeneity and identify prognostic LBD biomarkers.

A major strength of this study lies in its large, multicentre cohort, with more even representation of overlapping clinical diagnoses (PD, DLB, and iRBD), supported by detailed clinical assessment and established diagnostic criteria. This balanced representation enhances the generalisability of these transdiagnostic findings and enables evaluation of disease trajectories across all clinical LBD. The integration of longitudinal PD data provided insight into cognitive progression, while inclusion of iRBD allowed assessment of phenoconversion risk in prodromal disease. Identification of neural correlates for the spatiotemporal onset of visual hallucinations offers mechanistic insight into one of the most clinically disabling symptoms of LBD. Several limitations should be acknowledged. A key limitation of this study is the limited availability of biomarker confirmation across contributing cohorts. Diagnostic classification was primarily based on clinical criteria, as systematic assessment of α-synuclein SAA, CSF biomarkers, and other molecular measures was not available for all participants. Specifically, SAA availability remains limited in DLB cohorts and is not consistently observed across clinically well-characterised populations. Furthermore, biomarker-based exclusion of underlying neurodegenerative pathology in control participants was not uniformly available across cohorts and may have affected z-score normalisation and subtype boundaries. Analyses were restricted to T1-weighted MRI, which provides robust structural information but does not capture microstructural, metabolic or functional changes; future studies incorporating diffusion MRI, PET and functional modalities will offer complementary insights into network-level mechanisms, although it is considerably more challenging to acquire multiple modalities in such large patient samples. Subtype stability could not yet be evaluated owing to limited longitudinal follow-up in DLB, but this should be addressable through ongoing prospective data collection across harmonised multicentre cohorts. The naming convention used in this transdiagnostic study reflects regions of maximal subtype differentiation rather than the earliest detectable atrophy and does not preclude the involvement of other regions contributing meaningfully to variation in disease expression. Raw MRI data were unavailable for some datasets, necessitating the use of FreeSurfer-derived structural measures; however, all analyses were harmonised using standardised quality-control procedures and covariate adjustment to maximise comparability across sites. Phenoconversion analyses in iRBD were limited by small sample size, although consistent trends across subtypes support the biological plausibility of the observed trajectories, and they match recent biotypes in the literature. While we demonstrate the utility of transdiagnostic subtyping in predicting visual hallucinations, future work should incorporate more detailed and standardised clinical assessments^78,79^ to improve phenotypic precision and enhance clinical applicability for identifying and monitoring patients at risk. Finally, co-pathologies such as tau, amyloid, and TDP-43, which have been associated with dementia phenoconversion, were available for a subset of early-stage patients within the PPMI cohort and could not be explicitly modelled in iRBD, progressed PD, and DLB due to data unavailability. RBD assessment was heterogeneous across cohorts, combining polysomnography-confirmed diagnoses and questionnaire-based assessments. A proportion of participants were classified as Stage 0, reflecting individuals without sufficient or coherent atrophy patterns to be confidently aligned to a subtype trajectory. α-synuclein SAA positivity and DaT-SPECT binding ratios in Stage 0 did not differ significantly from those of classified subtype groups, indicating that pathology is present at broadly similar levels despite the absence of detectable atrophy. This group likely represents a heterogeneous mixture of early-stage cases with minimal structural involvement in whom functional or molecular changes may precede detectable atrophy, and potentially individuals with relative resilience or compensation delaying the emergence of measurable structural change. As primarily a structural imaging study, these individuals were excluded from subtype-specific analyses to minimise classification uncertainty. Future studies incorporating biomarker-enriched cohorts, genetics, and multimodal imaging will be essential to refine subtype boundaries and improve the biological interpretability of transdiagnostic models in LBD. Additionally, the prodromal spectrum in this study was enriched for iRBD, and other prodromal phenotypes (e.g., olfactory-predominant or MCI-LB) were underrepresented. Future studies should include these groups, as well as Alzheimer’s disease, to further refine subtype boundaries across the LBD–AD continuum.

In summary, this study adopts a transdiagnostic, biologically grounded framework for understanding heterogeneity across LBD, with implications for diagnosis, staging, and prediction. Imaging-derived atrophy subtypes reveal overlapping yet distinct trajectories that explain variation in cognition, core symptoms and phenoconversion risk. Conceptualising LBD as a continuum rather than as discrete clinical syndromes such as PD and DLB moves beyond traditional clinical distinctions, such as the ‘one-year rule’, and instead aligns classification with underlying mechanisms. By capturing the temporal sequence of atrophy progression, SuStaIn offers a more fine-grained index of disease stage than duration alone. Clinically, using such a framework has the potential to redefine the clinical diagnosis, identify conversion-prone individuals with iRBD, stratify patients with PD by risk of cognitive versus motor decline, and reduce heterogeneity in clinical trials through recruitment of biologically homogeneous cohorts. More broadly, it provides a data-driven approach that bridges pathology and clinical presentation, advancing efforts towards precision medicine in LBD and supporting a shift towards biological diagnostic frameworks.

## Contributors

All authors read and approved the final version of the manuscript. Author roles included: conception and design of the study (NPO, EM); collaboration and discussion (AK, GCL, NPO, EM), data acquisition (AK, NZ, AH, NCP, MCG, J-PT, MF, DA, AB, KS, FB, ATI, BS, CC-P, RW-M, MTH, RB, ID, ZW, DAR, EW, DF, SJGL, RSW, CL, NPO, EM); data analysis and figures (AK, GCL, EM), funding (RLR, CL, RSW, NPO, EM); writing – original draft (AK, EM); writing – review and final approval of manuscript (AK, GCL, NZ, AH, NCP, MCG, J-PT, MF, DA, AB, KS, FB, ATI, BS, CC-P, RW-M, MTH, RB, ID, ZW, DAR, EW, DF, GMH, SJGL, RSW, RLR, CL, NPO, EM). AK, GCL, NPO and EM directly accessed and verified the data reported in this manuscript.

## Data Availability

Data supporting the findings of this study are available from the corresponding author, upon reasonable request.

## Acknowledgements

We would like to acknowledge participants and caregivers for their efforts participating in this study.

AK was supported by The Australian Rotary Health/Rotary Club of Belconnen 50th Anniversary PhD Scholarship (SC4968).

GCL and NPO acknowledge funding from the UKRI Medical Research Council (MR/S03546X/1, MR/X024288/1).

EM was supported by a National Health and Medical Research Council (NHMRC) grant (2008565) and University of Sydney Horizon Fellowship.

MRI and clinical data from the Oxford Parkinson’s Disease Centre Discovery Cohort (OPDC) study, funded by the Monument Trust Discovery Award from Parkinson’s UK (J-1403), a charity registered in England and Wales (2581970) and in Scotland (SC037554), with the support of the National Institute for Health Research (NIHR) based at Oxford University Hospitals NHS Foundation Trust and the University of Oxford, and the NIHR Comprehensive Local Research Network.

MF and J-PT are supported by the NIHR Newcastle Biomedical Research Centre.

RSW is supported by a Wellcome Career Development Award (#225263/Z/22/Z), Rosetrees, Parkinson’s UK, the Lewy Body Society, Michael J Fox Foundation, and by the National Institute for Health and Care Research University College London Hospitals Biomedical Research Centre.

RB received support from the Wolfson Foundation and Eisai through a joint PhD fellowship.

CL was supported by the Medical Research Council (MR/R006504/1), Parkinson’s UK, Hilary-Galen Weston Foundation and Michael J Fox Foundation.

DA was supported by research grants from Institute of Health Carlos III (ISCIII), Spain, PI18/00435, PI22/00611, INT19/00016, INT23/00048 and PI25/00422 to DA, and by the Department of Health Generalitat de Catalunya PERIS programme SLT006/17/125, SLT042/25/000034.

ZW received funding from ARUK and Lewy Body Society. GMH was supported by a NHMRC Investigator grant (2034292).

RLR is supported by an NHMRC Emerging Leadership Grant (GNT2010064).

This research was funded in whole, or in part by the Wellcome Trust [227341/Z/23/Z].

The NACC database is funded by NIA/NIH Grant U24 AG072122. NACC data are contributed by the NIA-funded ADRCs: P30 AG062429 (PI James Brewer, MD, PhD), P30 AG066468 (PI Oscar Lopez, MD), P30 AG062421 (PI Teresa Gomez-Isla, MD), P30 AG066509 (PI Thomas Grabowski, MD), P30 AG066514 (PI Mary Sano, PhD), P30 AG066530 (PI Helena Chui, MD, Arthur Toga, PhD), P30 AG066507 (PI Marilyn Albert, PhD), P30 AG066444 (PI David Holtzman, MD), P30 AG066518 (PIs Lisa Silbert, MD, Kevin Duff, PhD), P30 AG066512 (PI Thomas Wisniewski, MD), P30 AG066462 (PI Scott Small, MD), P30 AG072979 (PI David Wolk, MD), P30 AG072972 (PIs Charles DeCarli, MD, Rachel Whitmer, PhD), P30 AG072976 (PI Andrew Saykin, PsyD), P30 AG072975 (PI Julie Schneider, MD, MS), P30 AG072978 (PI Ann McKee, MD), P30 AG072977 (PI Robert Vassar, PhD), P30 AG066519 (PI Joshua Grill, PhD), P30 AG062677 (PIs Brad Boeve, MD, Ronald Petersen, MD, PhD), P30 AG079280 (PI Jessica Langbaum, PhD), P30 AG062422 (PI Gil Rabinovici, MD), P30 AG066511 (PI Allan Levey, MD, PhD), P30 AG072946 (PI Linda Van Eldik, PhD), P30 AG062715 (PI Sanjay Asthana, MD, FRCP), P30 AG072973 (PI Russell Swerdlow, MD), P30 AG066506 (PIs Glenn Smith, PhD, ABPP, David Lowenstein, PhD, Ranjan Duara, MD), P30 AG066508 (PIs Stephen Strittmatter, MD, PhD, Christopher Van Dyck, MD), P30 AG066515 (PI Victor Henderson, MD, MS), P30 AG072947 (PI Suzanne Craft, PhD), P30 AG072931 (PI Henry Paulson, MD, PhD), P30 AG066546 (PIs Sudha Seshadri, MD, Gladys Maestre, MD, PhD), P30 AG086401 (PI Erik Roberson, MD, PhD), P30 AG086404 (PI Gary Rosenberg, MD), P30 AG086403 (PI Angela Jefferson, PhD), P30 AG072958 (PIs Heather Whitson, MD, Gwenn Garden, MD, PhD), P30 AG072959 (PI Jagan Pillai, MD, PhD), P30 AG092752 (Ihab Hajjar, MD, MS).

The NACC database is funded by NIA/NIH Grant U24 AG072122. SCAN is a multi-institutional project that was funded as a U24 grant (AG067418) by the National Institute on Aging in May 2020. Data collected by SCAN and shared by NACC are contributed by the NIA-funded ADRCs as follows: Arizona Alzheimer’s Center - P30 AG072980 (PI: Eric Reiman, MD); R01 AG069453 (PI: Eric Reiman (contact), MD); P30 AG019610 (PI: Eric Reiman, MD); and the State of Arizona which provided additional funding supporting our center; Boston University - P30 AG013846 (PI Neil Kowall MD); Cleveland ADRC - P30 AG062428 (James Leverenz, MD); Cleveland Clinic, Las Vegas – P20AG068053; Columbia - P50 AG008702 (PI Scott Small MD); Duke/UNC ADRC – P30 AG072958; Emory University - P30AG066511 (PI Levey Allan, MD, PhD); Indiana University - R01 AG19771 (PI Andrew Saykin, PsyD); P30 AG10133 (PI Andrew Saykin, PsyD); P30 AG072976 (PI Andrew Saykin, PsyD); R01 AG061788 (PI Shannon Risacher, PhD); R01 AG053993 (PI Yu-Chien Wu, MD, PhD); U01 AG057195 (PI Liana Apostolova, MD); U19 AG063911 (PI Bradley Boeve, MD); and the Indiana University Department of Radiology and Imaging Sciences; Johns Hopkins - P30 AG066507 (PI Marilyn Albert, Phd.); Mayo Clinic - P50 AG016574 (PI Ronald Petersen MD PhD); Mount Sinai - P30 AG066514 (PI Mary Sano, PhD); R01 AG054110 (PI Trey Hedden, PhD); R01 AG053509 (PI Trey Hedden, PhD); New York University - P30AG066512-01S2 (PI Thomas Wisniewski, MD); R01AG056031 (PI Ricardo Osorio, MD); R01AG056531 (PIs Ricardo Osorio, MD; Girardin Jean-Louis, PhD); Northwestern University - P30 AG013854 (PI Robert Vassar PhD); R01 AG045571 (PI Emily Rogalski, PhD); R56 AG045571, (PI Emily Rogalski, PhD); R01 AG067781, (PI Emily Rogalski, PhD); U19 AG073153, (PI Emily Rogalski, PhD); R01 DC008552, (M.-Marsel Mesulam, MD); R01 AG077444, (PIs M.-Marsel Mesulam, MD, Emily Rogalski, PhD); R01 NS075075 (PI Emily Rogalski, PhD); R01 AG056258 (PI Emily Rogalski, PhD); Oregon Health and Science University - P30 AG066518 (PIs Lisa Silbert, MD, Kevin Duff, PhD); R56 AG074321 (PI Jeffrey Kaye, MD); Rush University - P30 AG010161 (PI David Bennett MD); Stanford – P30AG066515; P50 AG047366 (PI Victor Henderson MD MS); University of Alabama, Birmingham – P20; University of California, Davis - P30 AG10129 (PI Charles DeCarli, MD); P30 AG072972 (PI Charles DeCarli, MD); University of California, Irvine - P50 AG016573 (PI Frank LaFerla PhD); University of California, San Diego - P30AG062429 (PI James Brewer, MD, PhD); University of California, San Francisco - P30 AG062422 (Rabinovici, Gil D., MD); University of Kansas - P30 AG035982 (Russell Swerdlow, MD); University of Kentucky - P30 AG028283-15S1 (PIs Linda Van Eldik, PhD and Brian Gold, PhD); University of Michigan ADRC - P30AG053760 (PI Henry Paulson, MD, PhD) P30AG072931 (PI Henry Paulson, MD, PhD) Cure Alzheimer’s Fund 200775 - (PI Henry Paulson, MD, PhD) U19 NS120384 (PI Charles DeCarli, MD, University of Michigan Site PI Henry Paulson, MD, PhD) R01 AG068338 (MPI Bruno Giordani, PhD, Carol Persad, PhD, Yi Murphey, PhD) S10OD026738-01 (PI Douglas Noll, PhD) R01 AG058724 (PI Benjamin Hampstead, PhD) R35 AG072262 (PI Benjamin Hampstead, PhD) W81XWH2110743 (PI Benjamin Hampstead, PhD) R01 AG073235 (PI Nancy Chiaravalloti, University of Michigan Site PI Benjamin Hampstead, PhD) 1I01RX001534 (PI Benjamin Hampstead, PhD) IRX001381 (PI Benjamin Hampstead, PhD); University of New Mexico - P20 AG068077 (Gary Rosenberg, MD); University of Pennsylvania - State of PA project 2019NF4100087335 (PI David Wolk, MD); Rooney Family Research Fund (PI David Wolk, MD); R01 AG055005 (PI David Wolk, MD); University of Pittsburgh - P50 AG005133 (PI Oscar Lopez MD); University of Southern California - P50 AG005142 (PI Helena Chui MD); University of Washington - P50 AG005136 (PI Thomas Grabowski MD); University of Wisconsin - P50 AG033514 (PI Sanjay Asthana MD FRCP); Vanderbilt University – P20 AG068082; Wake Forest - P30AG072947 (PI Suzanne Craft, PhD); Washington University, St. Louis - P01 AG03991 (PI John Morris MD); P01 AG026276 (PI John Morris MD); P20 MH071616 (PI Dan Marcus); P30 AG066444 (PI John Morris MD); P30 NS098577 (PI Dan Marcus); R01 AG021910 (PI Randy Buckner); R01 AG043434 (PI Catherine Roe); R01 EB009352 (PI Dan Marcus); UL1 TR000448 (PI Brad Evanoff); U24 RR021382 (PI Bruce Rosen); Avid Radiopharmaceuticals / Eli Lilly; Yale - P50 AG047270 (PI Stephen Strittmatter MD PhD); R01AG052560 (MPI: Christopher van Dyck, MD; Richard Carson, PhD); R01AG062276 (PI: Christopher van Dyck, MD); 1Florida - P30AG066506-03 (PI Glenn Smith, PhD); P50 AG047266 (PI Todd Golde MD PhD).

PPMI – a public-private partnership – is funded by the Michael J. Fox Foundation for Parkinson’s Research and funding partners, including AbbVie, Alamar Biosciences, Aligning Science Across Parkinson’s (ASAP), Arrowhead Pharma, Arvinas, AskBio, BIAL, BioArctic, Biohaven, BlueRock Therapeutics, Bristol Myers Squibb, Calico Labs, Capsida Biotherapeutics, Critical Path Institute, DaCapo Brainscience, Denali, Edmond J. Safra Foundation, Eli Lilly, Gain Therapeutics, GE Healthcare, Genentech, GSK, Insitro, Johnson & Johnson Innovative Medicine, Lundbeck, Merck, Neumora, Neuron23, Novartis, Olink, Regeneron, Roche, Sanofi, Tenvie, UCB, Vanqua Bio, Voyager Therapeutics, The Weston Family Foundation.

## Data sharing statement

Data supporting the findings of this study are available from the corresponding authors, upon reasonable request. pySuStaIn algorithm can be publicly accessed at https://github.com/ucl-pond/pySuStaIn.

