## Supplementary Materials for "Data-driven trajectories of atrophy explain clinical heterogeneity across Lewy body diseases"

**Supplementary Methods**

**CSF Biomarker Analysis**

Cerebrospinal fluid (CSF) collection, handling, shipment, and storage were conducted according to standardized operating procedures at each participating site, as detailed in the PPMI biologics manual. Samples were processed and distributed through the PPMI Biorepository Core Laboratories and subsequently analysed at the University of Pennsylvania Biomarker Research Laboratory. Core Alzheimer’s disease biomarkers, including amyloid-β₁₋₄₂, total tau (t-tau), and phosphorylated tau at threonine 181 (p-tau181), were quantified using Elecsys electrochemiluminescence immunoassays on the cobas e601 platform (Roche Diagnostics), following established protocols.

**Supplementary Results**

**Striatal dopaminergic vulnerability of early PD across transdiagnostic subtypes**

DaT-SPECT revealed differential dopaminergic vulnerability across classified subtypes. Subtypes A and B both demonstrated significant annual dopaminergic decline across all striatal subregions. The strongest effects were observed in the left caudate for subtype A (β = −0.057 SBR/year, 95% CI [−0.081, −0.033], p_FDR_ < 0.001) and left striatum for subtype B (β = −0.035 SBR/year, 95% CI [−0.056, −0.014], p_FDR_ = 0.002), with rates of loss that did not significantly differ between A and B (all p_FDR_ > 0.05). Subtype C showed no significant annual change in dopaminergic binding across any striatal subregion (all p > 0.05), and was significantly preserved relative to Subtypes A and B in the left striatum (A: β = −0.048, 95% CI [−0.069, −0.027], p_FDR_ = 0.025; B: β = −0.035, 95% CI [−0.056, −0.014], p_FDR_ = 0.040) and left caudate (A: β = −0.057, 95% CI [−0.081, −0.033], p_FDR_ = 0.019; B: β = −0.038, 95% CI [−0.063, −0.014], p_FDR_ = 0.056), confirming preserved striatal integrity. Subtype D has limited interpretability due to limited sample available (n = 2).

**Characterisation of subtypes using CSF biomarkers**

To characterise the biological underpinnings of the identified LBD subtypes, we examined CSF biomarkers of AD co-pathology (Aβ_1-42_, pTau, tTau, NfL, and the pTau/Aβ_1-42_ ratio), α-synuclein SAA positivity that was available for a subset of early-stage individuals with PD from the PPMI cohort (Supplementary Table S4). α-synuclein SAA positivity did not differ between stage 0 patients with PD and classified subtypes; however, subtype C (Early temporo-limbic/late basal ganglia; 100%) had significantly higher positivity than Subtype D (Early basal ganglia–cingulate/late cortex; 50%) (p_FDR_ = 0.039). AD biomarker profiles were broadly similar across subtypes and were not significantly different. This lack of differences could be attributed to limited sample size and the early disease stage of the cohort, which may reduce sensitivity to detect subtype-related differences in AD co-pathology.

**RBD prevalence varies across LBD subtypes**

We examined stage-related changes in RBD prevalence within clinically diagnosed PD and DLB using multivariable logistic regression adjusted for age and sex (Supplementary Figure S2A). Stage was not associated with RBD prevalence in either diagnosis. In disease-stratified analyses, however, DLB subtype D showed higher RBD rates than subtype B (pFDR < 0.031; χ² = 7.771) (Supplementary Figure S2B); whereas no subtype differences were detected within PD.

**Clinical profiles of diagnosis-specific models**

Subtype-specific analyses within PD and DLB showed expected clinical differences (Supplementary Tables S2 and S3, Figure S6). These findings confirm that SuStaIn recovers biologically plausible atrophy trajectories within individual diagnostic groups. Within DLB, DLB-A subtype had fewer years of education than DLB-C (p_FDR_=0.045) and significantly higher age and lower disease duration than the DLB-B (p_FDR_=0.009) subtype. Within PD, the PD-C subtype was older than both PD-A (p_FDR_<0.001) and PD-B (p=0.049). The PD-C group also had longer disease duration since diagnosis compared to the PD-A and PD-B subtypes (all p_FDR_<0.001). Likewise, the PD-C group exhibited more severe motor symptoms (MDS-UPDRS-III) than the PD-A (p_FDR_<0.001) and PD-B groups (p_FDR_<0.001). Patients in the PD-B subtype had significantly higher depression scores relative to the PD-A subtype (p<0.001) and higher Sniffin’ sticks scores than the subcortical subtype (p=0.011).

**Tables:**

- Supplementary Table S1: Site-wise distribution of clinical diagnoses across SuStaIn subtypes
- Supplementary Table S2: Feature Selection for SuStaIn modelling
- Supplementary Table S3: Demographics by clinical diagnosis
- Supplementary Table S4. CSF biomarker profiles and α-synuclein seed amplification assay positivity across PD subtypes.
- Supplementary Table S5: Demographics of patients with PD used for longitudinal analyses.
- Supplementary Table S6: Demographics of iRBD-PD and iRBD-DLB converters.
- Supplementary Table S7: Demographic information of classified and unclassified DLB subtyped using the DLB-only model
- Supplementary Table S8: Demographic information of classified and unclassified PD subtyped using the PD-only model.

**Figures:**

- Supplementary Figure S1: Criteria used to select the optimal number of subtypes (combined and disease-specific)
- Supplementary Figure S2: Subtype assignment probability across SuStaIn stage
- Supplementary Figure S3: RBD prevalence by SuStaIn stage
- Supplementary Figure S4: Regional atrophy and stage-dependent emergence of visual hallucinations across subtypes
- Supplementary Figure S5: Baseline subtype in iRBD individuals predicts phenoconversion.
- Supplementary Figure S6: Spatiotemporal atrophy signatures of DLB subtypes
- Supplementary Figure S7: Spatiotemporal atrophy signatures of PD subtypes
- Supplementary Figure S8: Clinical profiles by disease specific subtypes in PD and DLB

| **Centre/Cohort** | **Group** | **Unclassified (n=123)** | **Early cortico-limbic/ late basal ganglia (A, n = 297)** | **Early basal ganglia/ late limbic (B, n=215)** | **Early temporo-limbic/late basal ganglia (C, n=123)** | ***Early basal ganglia***–***cingulate/late cortex (D, n=75)*** | **Total** |
| --- | --- | --- | --- | --- | --- | --- | --- |
| Newcastle | PD | 2 | 7 | 1 | 11 | 1 | 22 |
|  | DLB | 8 | 64 | 13 | 3 | 2 | 90 |
|  | iRBD | 0 | 0 | 0 | 0 | 1 | 1 |
| Sydney | PD | 6 | 17 | 46 | 13 | 20 | 102 |
|  | DLB | 0 | 6 | 0 | 1 | 9 | 16 |
|  | iRBD | 5 | 11 | 4 | 1 | 25 | 46 |
| OPDC | PD | 17 | 37 | 27 | 2 | 2 | 85 |
|  | iRBD | 9 | 23 | 3 | 35 | 1 | 71 |
| VIP | PD | 1 | 2 | 6 | 2 | 0 | 11 |
|  | DLB | 7 | 11 | 5 | 16 | 1 | 40 |
| Brugman | DLB | 1 | 22 | 7 | 0 | 0 | 30 |
|  | iRBD | 0 | 1 | 0 | 0 | 1 | 2 |
| Izmir | DLB | 2 | 16 | 3 | 2 | 1 | 24 |
|  | iRBD | 1 | 1 | 0 | 0 | 0 | 2 |
| Istanbul | DLB | 2 | 3 | 0 | 1 | 0 | 6 |
| LAFE | DLB | 1 | 3 | 1 | 0 | 2 | 7 |
| Santpau | DLB | 6 | 21 | 1 | 3 | 2 | 33 |
| NACC | DLB | 6 | 14 | 1 | 5 | 3 | 29 |
| PPMI | PD | 35 | 36 | 49 | 21 | 4 | 145 |
| qMAP-PD | PD | 14 | 2 | 48 | 7 | 0 | 71 |

**Supplementary Table S1: Site-wise distribution of clinical diagnoses across SuStaIn subtypes.** Counts of participants by centre and diagnosis (PD, DLB, iRBD) allocated to stage 0 and the four data-driven subtypes at baseline. Subtype labels correspond to the combined-model trajectories: A (Early cortico-limbic/late basal ganglia), B (Early basal ganglia/ late limbic), C (Early temporo-limbic/late basal ganglia), and D (Early basal ganglia–cingulate/late cortex). PD, Parkinson’s disease; DLB, dementia with Lewy bodies; iRBD, idiopathic REM sleep behaviour disorder.

| **Lobe** | **Region of Interest** | **PCA Contribution** |
| --- | --- | --- |
| Parietal Lobe | rh_precuneus_volume | 0.152474955 |
|  | lh_precuneus_volume | 0.140393617 |
|  | rh_inferiorparietal_volume | 0.128720362 |
|  | lh_supramarginal_volume | 0.122009394 |
|  | lh_inferiorparietal_volume | 0.120074574 |
|  | rh_supramarginal_volume | 0.119310395 |
|  | lh_superiorparietal_volume | 0.099863599 |
|  | rh_superiorparietal_volume | 0.098130822 |
| Frontal Lobe | lh_lateralorbitofrontal_volume | 0.147873389 |
|  | lh_superiorfrontal_volume | 0.138134943 |
|  | rh_medialorbitofrontal_volume | 0.136681036 |
|  | rh_lateralorbitofrontal_volume | 0.135599798 |
|  | rh_superiorfrontal_volume | 0.130442404 |
|  | lh_medialorbitofrontal_volume | 0.129469713 |
|  | lh_rostralmiddlefrontal_volume | 0.1132002 |
|  | rh_rostralmiddlefrontal_volume | 0.111507472 |
|  | lh_caudalmiddlefrontal_volume | 0.105752254 |
|  | rh_caudalmiddlefrontal_volume | 0.090728395 |
|  | rh_frontalpole_volume | 0.079744124 |
|  | lh_frontalpole_volume | 0.076153555 |
| Occipital Lobe | lh_fusiform_volume | 0.141849392 |
|  | rh_fusiform_volume | 0.137425137 |
|  | lh_lateraloccipital_volume | 0.123428214 |
|  | rh_lateraloccipital_volume | 0.120455016 |
|  | rh_lingual_volume | 0.109528663 |
|  | lh_lingual_volume | 0.108594474 |
|  | rh_cuneus_volume | 0.091380254 |
|  | lh_pericalcarine_volume | 0.090979062 |
|  | rh_pericalcarine_volume | 0.085190675 |
|  | lh_cuneus_volume | 0.080999829 |
| Temporal Lobe | rh_superiortemporal_volume | 0.130155065 |
|  | lh_superiortemporal_volume | 0.130105613 |
|  | rh_middletemporal_volume | 0.12829738 |
|  | lh_middletemporal_volume | 0.121699246 |
|  | rh_transversetemporal_volume | 0.120633027 |
|  | rh_inferiortemporal_volume | 0.119506933 |
|  | lh_inferiortemporal_volume | 0.119368969 |
|  | lh_transversetemporal_volume | 0.118082528 |
|  | lh_temporalpole_volume | 0.10782703 |
|  | rh_temporalpole_volume | 0.106825391 |
| Insula | lh_insula_volume | 0.148759273 |
|  | rh_insula_volume | 0.137031965 |
| Precentral | lh_precentral_volume | 0.140487142 |
|  | rh_precentral_volume | 0.13489316 |
| Posterior Cingulate | lh_posteriorcingulate_volume | 0.112546243 |
|  | rh_posteriorcingulate_volume | 0.108977492 |
| Parahippocampus | rh_parahippocampal_volume | 0.102793969 |
|  | lh_parahippocampal_volume | 0.089916116 |
| Anterior Cingulate | lh_rostralanteriorcingulate_volume | 0.099389813 |
|  | rh_rostralanteriorcingulate_volume | 0.08398694 |
|  | rh_caudalanteriorcingulate_volume | 0.067121364 |
|  | lh_caudalanteriorcingulate_volume | 0.057166241 |
| Hippocampus | Right-Hippocampus | 0.095946001 |
|  | Left-Hippocampus | 0.094495721 |
| Putamen | Right-Putamen | 0.092317089 |
|  | Left-Putamen | 0.087438437 |
| Amygdala | Left-Amygdala | 0.085478562 |
|  | Right-Amygdala | 0.084691826 |
| Entorhinal | rh_entorhinal_volume | 0.081973071 |
|  | lh_entorhinal_volume | 0.076819579 |
| Thalamus | Right-Thalamus | 0.077053236 |
|  | Left-Thalamus | 0.057598671 |
| Caudate | Right-Caudate | 0.05635302 |
|  | Left-Caudate | 0.049356134 |
| Accumbens* | Right-Accumbens-area | 0.054545351 |
|  | Left-Accumbens-area | 0.040323554 |
| Pallidum* | Right-Pallidum | 0.045839067 |
|  | Left-Pallidum | 0.043321097 |
| Brain-Stem* | Brain-Stem | 0.041906188 |

**Supplementary Table S2: Feature Selection for SuStaIn modelling.** Final set of ROIs used for PD-only, DLB-only and LBD models along with their 1^st^ component loadings. ROIs were selected using a hybrid method both PCA and literature-based findings. * indicate ROIs selected based on literature. PD, Parkinson’s disease; DLB, dementia with Lewy bodies; iRBD, idiopathic REM sleep behaviour disorder.

| **Clinical Variables** | **HC (*n*=452)** | **PD (*n*=436)** | | **DLB (n=275)** | | **iRBD (*n*=122)** |
| --- | --- | --- | --- | --- | --- | --- |
| Age* | 65 [64, 66]^ | 65.15 [64.36, 66]^d^ | | 76 [75, 77]^^,d,f^ | | 66.52 [64.68, 68]^f^ |
| Sex* | 251:215^a,b,c^ | 284:152^a,e^ | | 191:84^b,f^ | | 109:13^c^ |
| Education* | 14 [13, 15]  (n = 380) | 15 [15, 16]  (n=357) | | 11 [11, 12]^d,f^  (n=269) | | 15 [15, 15]^f^  (n=108) |
| Disease Duration (years)* | - | 1.76 [1.52, 2.03]^d,e^ (n=380) | | 1.25 [0.92, 1.71]^d^  (n=156) | | 1.20 [0.93, 1.42]^e^ (n=113) |
| **Questionnaires** | | |  | |  | |
| Geriatric Depression Scale* | 0 [0, 0]^a^  (n=216) | 5 [5, 5]^a^  (n=214) | | - | | - |
| RBD Screening Questionnaire* | 2 [2, 3]^a,b,c^  (n=277) | 3 [3, 4]^a,d^  (n=306) | | 8 [7, 9]^b,d,e,f^  (n=40) | | 10 [10, 11]^c,e,f^  (n=67) |
| HADS Anxiety* | - | 4 [3, 4]^f^  (n=89) | | 5.5 [4, 7]  (n=38) | | 3 [2, 4]^f^  (n=104) |
| HADS Depression* | - | 3 [2, 4]^d,e^  (n=89) | | 5 [4.5, 6.5]^d,f^  (n=38) | | 2 [1, 3]^e,f^  (n=104) |
| **Cognitive Measures** | | |  | |  | |
| Montreal Cognitive Assessment* | 28 [28, 28]^a,b,c^  (n=240) | 27 [27, 28]^a,d^  (n=373) | | 20 [19, 22] ^b,d,f^  (n=88) | | 27 [26, 28]^c,f^  (n=41) |
| Mini Mental State Examination* | 29 [29, 29]^a,b,c^  (n=205) | 29 [28, 29]^a,d^  (n=193) | | 25 [24, 25.5]^b,d,f^  (n=228) | | 28 [28, 28]^c,f^  (n=68) |
| **Sensory/Motor** | | |  | |  | |
| Sniffin Sticks* | - | 7 [6, 8]^e^  (n=110) | | - | | 8 [7, 8]^e^  (n=95) |
| MDS-UPDRS III* | 0 [0, 1]^a,b,c^  (n=339) | 22 [20.5, 24]^a,e^  (n=416) | | 20 [17, 22]^b,f^  (n=198) | | 5 [3.5, 6.5]^c,e,f^  (n=106) |
| **Stage 0 (n)** | - | 75 | | 33 | | 15 |

**Supplementary Table S3: Demographics by clinical diagnosis.** Median [95% CI] of demographic and clinical data across the healthy controls, Parkinson’s disease, dementia with Lewy bodies and isolated rapid eye movement behaviour disorder. Clinical measures include: GDS (Geriatric Depression Scale), HADS (Hospital anxiety and depression scale), MDS-UPDRS–III (Movement Disorder Society Unified Parkinson’s Disease Rating Scale Section III), MMSE (Mini Mental State Examination), MoCA (Montreal Cognitive Assessment), RBDSQ (Rapid eye movement behaviour disorder screening questionnaire). Pairwise post-hoc comparisons (FDR corrected, p<0.05) are denoted as: * indicates significant differences between groups. a (HC vs PD), b ^(^HC vs DLB), c (HC vs iRBD), d ^(^PD vs DLB), e (PD vs iRBD), f (DLB vs iRBD)**.** Age, sex and stage are available for all individuals.

| **Clinical Variables** | **Stage 0** | **Early cortico-limbic/late basal ganglia** | **Early basal ganglia/late limbic** | **Early temporo-limbic/late basal ganglia** | **Early basal ganglia–cingulate/late cortex** |
| --- | --- | --- | --- | --- | --- |
| **Aβ_1-42_ (n)** | 1031.2±526.3  (n=20) | 944.4±416.0  (n=22) | 1011.3±365.4 (n=35) | 762.6 ± 223.9 (n=14) | 985.8±718.01  (n=4) |
| **pTau (n)** | 15.6±7.6  (n=25) | 15.4±5.9  (n=25) | 16.7±8.7  (n=36) | 10.7±1.9  (n=18) | 15.3±8.0  (n=4) |
| **tTau (n)** | 179.9±78.2  (n=25) | 178.7±62.5  (n=25) | 192.9±88.3 (n=36) | 135.0±36.3 (n=18) | 176.9±87.9  (n=4) |
| **NfL (n)** | 14.6 ± 7.3  (n=24) | 26.8± 41.6  (n=27) | 12.6±5.7  (n=35) | 16.0±10.6  (n=16) | 12.7±5.5  (n=4) |
| **pTau/Aβ_1-42_ Ratio (n)** | 0.0184 ± 0.0128  (n=20) | 0.0188 ± 0.0107  (n=22) | 0.0195 ± 0.0184  (n=35) | 0.0146 ± 0.0032  (n=14) | 0.0182 ± 0.0092  (n=4) |

**Supplementary Table S4. CSF biomarker profiles and α-synuclein seed amplification assay positivity across PD subtypes.** Values for biomarkers are presented as mean ± SD with sample size in parentheses. αSyn SAA positivity is expressed as percentage of positive cases with sample size in parentheses. Aβ_1-42_, Amyloid beta_1-42_; pTau, phosphorylated tau; tTau, total tau; NfL, neurofilament light chain; pTau/Aβ_1-42_ ratio, phosphorylated tau to amyloid beta_1-42_ ratio; αSyn SAA, alpha-synuclein seed amplification assay; CSF, cerebrospinal fluid.

| **Clinical Variables** | **Early cortico-limbic/late basal ganglia**  **(A, n = 39)** | **Early basal ganglia/late limbic**  **(B, n = 63)** | | **Early temporo-limbic/late basal ganglia**  **(C, n = 24)** | | **Early basal ganglia**–**cingulate/late cortex**  **(D, n = 11)** |
| --- | --- | --- | --- | --- | --- | --- |
| Age* | 61.8 [59.6, 64.7]^a,c^  (n=39) | 68.2 [65.1, 71.4]^a^  (n=63) | | 62.9 [54.4, 69.3]  (n=24) | | 68.4 [66.0, 75.0]^c^  (n=11) |
| Sex | 18:21 | 26:37 | | 13:11 | | 8:3 |
| Education | 14 [13, 16]  (n=39) | 16 [16, 16]  (n=63) | | 16 [15, 18]  (n=24) | | 13 [10, 17]  (n=11) |
| Disease Duration (years)* | 1.25 [0.83, 1.66]  (n = 39)^c^ | 1 [0.5, 1.75]  (n = 63)^e^ | | 2.08 [0.83, 3.16]  (n = 24) | | 6.33 [1.91, 8.66]  (n = 11)^c,e^ |
| Follow-up (months) | 26 [13.2, 37.2]  (n = 39) | 39.6 [24, 60]  (n=63) | | 42 [13, 71]  (n=24) | | 72 [42, 101]  (n=11) |
| MCI converters | 22/39^a^ | 19/63^a^ | | 11/24 | | 6/11 |
| **Questionnaires** | | |  | |  | |
| RBD Screening Questionnaire | 6 [5, 8.5]  (n=12) | 6 [5, 9]  (n=17) | | 6.5 [5, 9.5]  (n=6) | | - |
| **Cognitive Measures** | | |  | |  | |
| Montreal Cognitive Assessment* | 26 [25, 28]  (n=39) | 28 [27, 29]  (n=63) | | 27.5 [25, 29]  (n=24) | | 25 [22, 28]  (n=11) |
| **Sensory/Motor** | | |  | |  | |
| MDS-UPDRS III* | 22 [16, 23.5]^c^  (n=38) | 21 [19, 25]^e^  (n=59) | | 17 [14, 25]^f^  (n=23) | | 36 [19, 51]^c,e,f^  (n=11) |

**Supplementary Table S5: Demographics of patients with PD used for longitudinal analyses. A–D** (stage 0 excluded; subtype probability ≥ 0.50). Values are in median [95% CI] unless stated. Letter markers denote pairwise comparisons among classified subtypes: a (A vs B), b (B vs C), c (A vs D), d (B vs C), e (B vs D),f (C vs D) MoCA, Montreal Cognitive Assessment; MDS-UPDRS III, Movement Disorder Society Unified Parkinson’s Disease Rating Scale part III; RBDSQ, REM Sleep Behavior Disorder Screening Questionnaire; PD, Parkinson’s disease.

| **Clinical Variables** | **Non-converters**  **(n=24)** | **PD converters**  **(n=10)** | | **DLB converters**  **(n=7)** |
| --- | --- | --- | --- | --- |
| Age | 66 [62, 68]  (n=24) | 65[62, 72]  (n=10) | | 74 [64, 77]  (n=7) |
| Sex (M:F) | 21:6 | 9:1 | | 7:0 |
| Education | 15 [13, 17]  (n=24) | 12.5 [10, 16]  (n=10) | | 12 [10, 13]  (n=7) |
| Disease duration (years) | 0.75 [0.33, 1.83]  (n=24) | 1.12 [0.75, 1.66]  (n=10) | | 3.08 [0.5, 4.33]  (n=7) |
| Time to conversion (years) | - | 2.54 [0.66, 5.58]  (n = 10) | | 1.50 [1.00, 4.41]  (n = 7) |
| **Questionnaires** |  |  | |  |
| RBD Screening Questionnaire | 5.0 [5.0, 9.0] | 11.0 [7.0, 12.0] | | 9.0 [5.0, 11.0] |
| **Cognitive Measures** |  |  | |  |
| Montreal Cognitive Assessment | 28 [27, 29]^a^  (n=24) | 28 [26, 29]^b^  (n=8) | | 23.5 [21, 25]^a,b^  (n=6) |
| **Sensory/Motor** | | |  | |
| MDS-UPDRS III | 8 [4, 11]  (n=23) | 7 [1, 22]  (n=9) | | 16 [0, 29]  (n=5) |
| **Stage** | 4 [3, 6] | 2.5 [1, 9.5] | | 4 [1,14] |

**Supplementary Table S6: Demographics of iRBD-PD and iRBD-DLB converters.** Median [95% CI] of demographic and clinical data between PD and DLB converters. Clinical measures include MDS-UPDRS–III (Movement Disorder Society Unified Parkinson’s Disease Rating Scale Section III), MoCA (Montreal Cognitive Assessment), RBDSQ (Rapid eye movement behaviour disorder screening questionnaire), Group differences were checked using non parametric permutation tests (FDR corrected, p < 0.05) are denoted as: * indicates significant differences between groups. Letter markers denote pairwise comparisons among classified subtypes: a (Non-converters vs PD converters), b (Non converters vs DLB converters)

| **Clinical Variables** | **Stage 0 (n=38)** | **DLB-A (n=81)** | | **DLB-B (n=118)** | | **DLB-C (n=37)** |
| --- | --- | --- | --- | --- | --- | --- |
| Age* | 76.5 [58.85, 83.07] | 77[64, 90]^a^ | | 75[61.92, 85.15]^a^ | | 74[61.5, 85.2] |
| Sex | 30:8 | 51:30 | | 84:34 | | 25:12 |
| Education* | 14 [12, 17]  (n=38) | 10 [9, 10]^b^  (n=79) | | 11 [10, 12]  (n=116) | | 12 [7, 13]^b^  (n=33) |
| Disease Duration (years)* | 3.5 [1.42, 6.08]  (n=14) | 1.17 [0, 1.58]^a^  (n=25) | | 2.54 [1.62, 3.08]^a^  (n=54) | | 2.38 [0.58, 3.5]  (n=24) |
| **Cognitive Measures** | | |  | |  | |
| Mini Mental State Examination | 25 [24, 27]  (n=34) | 23 [21, 26]  (n=70) | | 24.5 [23, 26]  (n=92) | | 25 [22, 26]  (n=32) |
| **Sensory/Motor** | | |  | |  | |
| MDS-UPDRS III | 19 [12, 23]  (n=28) | 19 [15, 23]  (n=55) | | 18 [14, 22]  (n=87) | | 30 [22.5, 38]  (n=28) |
| **Stage** | 0 | 18 [1, 38] | | 14 [1, 37.5] | | 5 [1, 28.1] |

**Supplementary Table S7: Demographic information of classified and unclassified DLB subtyped using the DLB-only model.** Median [95% CI] of demographic and clinical data across the three classified DLB subtypes and the unclassified group. Clinical measures include: MDS-UPDRS–III (Movement Disorder Society Unified Parkinson’s Disease Rating Scale Section III), MMSE (Mini Mental State Examination). Pairwise post-hoc comparisons (FDR corrected, p<0.05) are denoted as: * indicates significant differences between groups a (DLB-A vs DLB-B), b (DLB-A vs DLB-C). Age, sex and stage are available for all individuals.

| **Clinical Variables** | **Stage 0 (n=69)** | **PD-A**  **(n=121)** | | **PD-B**  **(n=125)** | | **PD-C**  **(n=104)** |
| --- | --- | --- | --- | --- | --- | --- |
| Age* | 65.4 [47.8, 78.17] | 62.1[39, 79]^a,c^ | | 65[47, 80.15]^a,b^ | | 67[50.58, 80.08]^b,c^ |
| Sex | 46:23 | 71:50 | | 82:43 | | 72:32 |
| Education* | 14.5 [13, 15.5]  (n=60) | 15 [14, 16]  (n=105) | | 16 [16, 17]^*^  (n=77) | | 15 [15, 17]^*^  (n=98) |
| Disease Duration (years)* | 1.14 [0.61, 1.52]^^^  (n=60) | 1.52 [1.19, 3.32]^c^  (n=97) | | 1.32 [1.05, 1.65]^b^ (n=114) | | 5.67 [4.54, 7]^^,b,c^  (n=98) |
| **Questionnaires** |  |  | |  | |  |
| Geriatric Depression Scale | 5 [5, 5]  (n=44) | 5 [4, 5]  (n=63) | | 4 [3, 5.5]  (n=90) | | 4 [3, 4]  (n=11) |
| RBD Screening Questionnaire^ | 4 [3, 5]^^^  (n=63) | 4 [4, 5]  (n=37) | | 3 [2, 4]^^^  (n=112) | | 3 [0, 3]  (n=15) |
| HADS Anxiety | 4 [1, 6]  (n=19) | 4 [2, 5]  (n=37) | | 3 [2, 4]  (n=24) | | 2 [0, 4]  (n=5) |
| **Cognitive Measures** |  |  | |  | |  |
| Mini Mental State Examination | 28 [27, 29.5]  (n=22) | 29 [29, 30]  (n=51) | | 28[27, 28]  (n=32) | | 29 [29, 30]  (n=76) |
| Trail Making Task-B | 66 [29, 71]  (n=15) | 64 [48.1, 72]  (n=21) | | 68 [59, 76]  (n=63) | | 89 [75, 106]  (n=75) |
| **Sensory/Motor** | | |  | |  | |
| MDS-UPDRS III* | 24 [20, 27]  (n=69) | 20.5 [18, 24]^c^  (n=118) | | 18.5 [17, 20.5]^b^  (n=120) | | 26 [22, 28]^b,c^  (n=93) |
| Sniffin’ Sticks^* | 9 [8, 11]^^^  (n=19) | 7 [5, 8]^c^  (n=37) | | 6 [5, 7]  (n=26) | | 4.5[1.5, 9]^^,c^  (n=28) |
| **Stage** | 0 | 3 [1, 15] | | 6 [1, 21.8] | | 14 [2, 30.42] |

**Supplementary Table S8: Demographic information of classified and unclassified PD subtyped using the PD-only model.** Median [95% CI] of demographic and clinical data across the three classified PD subtypes and the unclassified group. Clinical measures include: GDS (Geriatric Depression Scale) HADS A&D (Hospital Anxiety and Depression scale), MDS-UPDRS–III (Movement Disorder Society Unified Parkinson’s Disease Rating Scale Section III), MMSE (Mini Mental State Examination), RBDSQ (Rapid eye movement behaviour disorder screening questionnaire), TMT-B (Trail Making Test Section B), Sniffin’ Sticks. Pairwise post-hoc comparisons (FDR corrected, p<0.05) are denoted as: * indicates significant differences between classified subtypes; ^ indicates differences between the unclassified and classified subtypes. Letter markers denote pairwise comparisons among classified subtypes: a (PD-A vs PD-B), b (PD-A vs PD-C), c (PD-B vs PD-C). Age, sex and stage are available for all individuals.

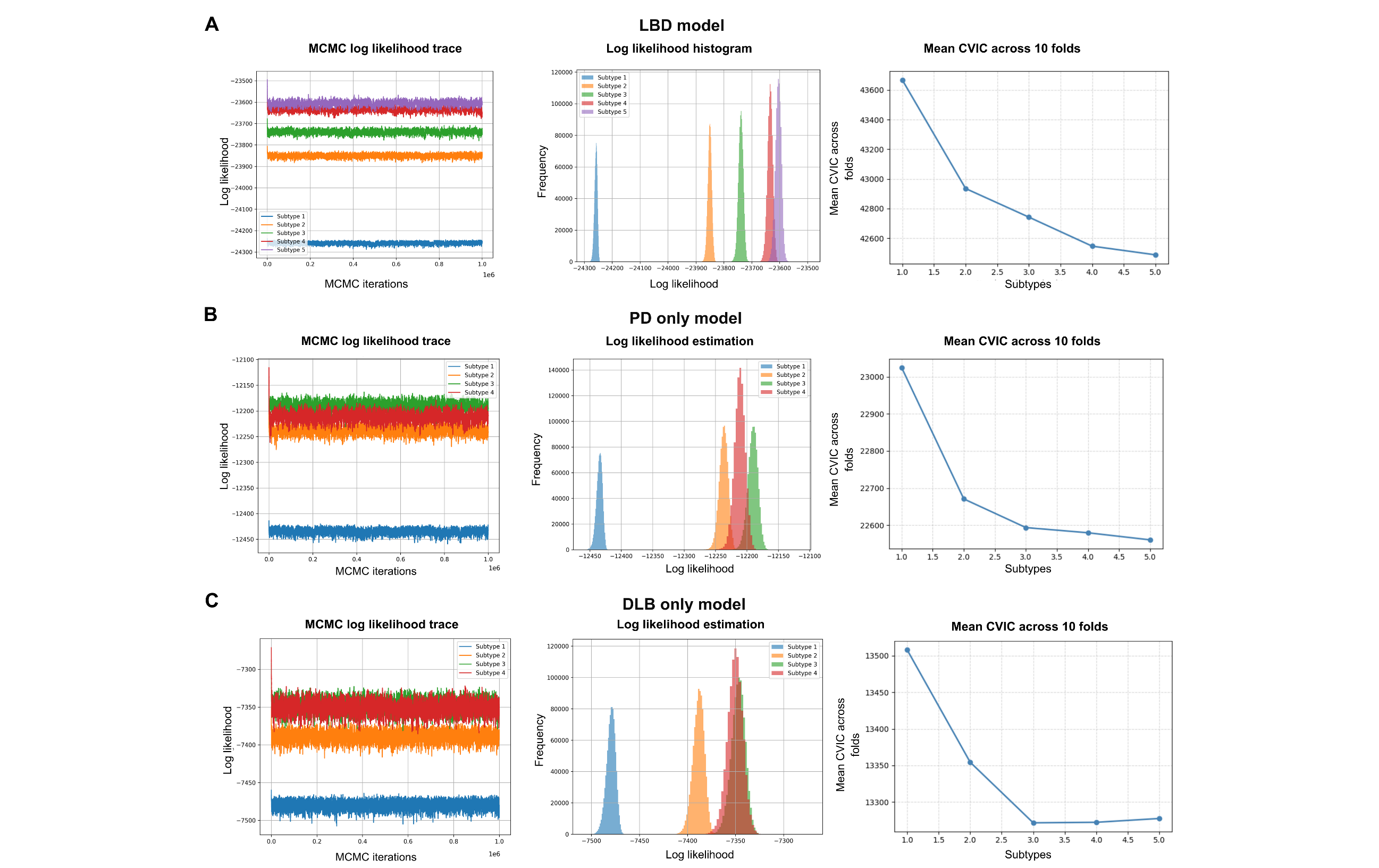

**Supplementary Figure S1: Criteria used to select the optimal number of subtypes (combined and disease-specific).** Cross-validation and Markov chain Monte Carlo (MCMC) diagnostics support a four-subtype solution for the combined LBD model and for PD-only and DLB-only analyses. **A, D, G:** MCMC log-likelihood traces by candidate subtype number; **B, E, H** corresponding log-likelihood histograms; **C, F, I** mean cross-validation information criterion (CVIC) across 10 folds versus number of subtypes (lower is better). **A–C,** Combined LBD model: **D–F,** PD-only model: **G–I,** DLB-only model. LBD (PD + DLB + iRBD model); PD, Parkinson’s disease; DLB, dementia with Lewy bodies.

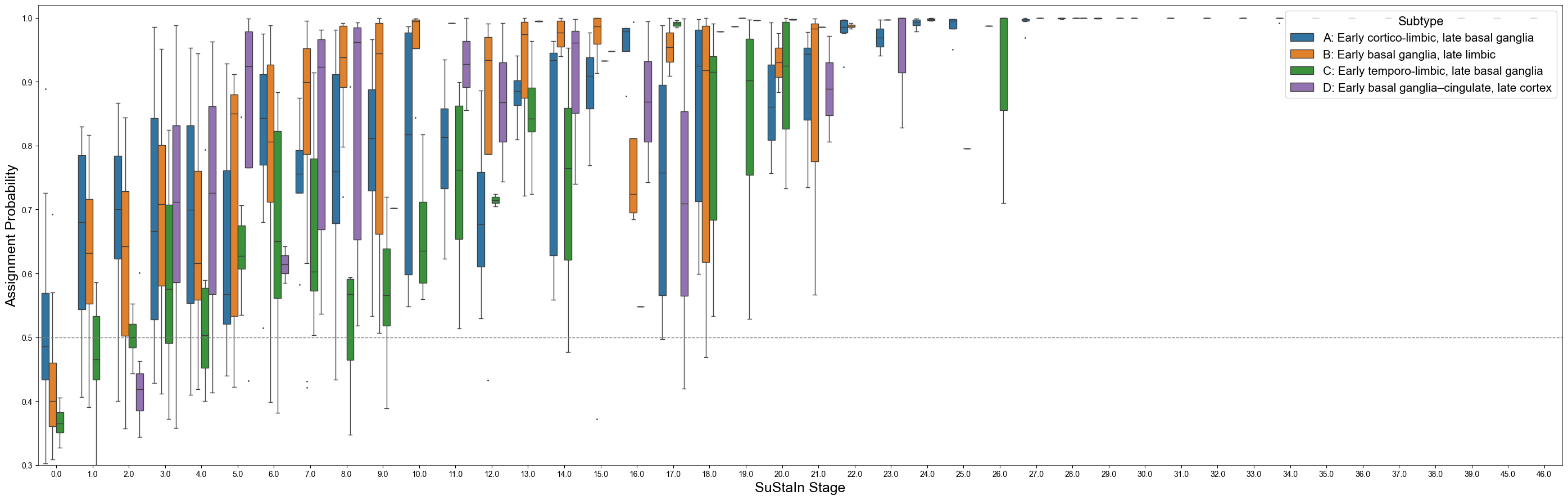

**Supplementary Figure S2: Subtype Probability assignment for each subtype across SuStaIn stages.** Boxplots display the distribution of assignment probabilities for each subject’s most likely subtype across SuStaIn stages (x-axis) across all subtypes. Each point reflects an individual’s probability of belonging to their assigned subtype, with 1.0 indicating complete certainty. Blue = Subtype A (Early cortico-limbic/late basal ganglia), Orange = Subtype B (Early basal ganglia/late limbic), Green = Subtype C (Early temporo-limbic/late basal ganglia), Purple = Subtype D (Early basal ganglia-cingulate/late cortex).

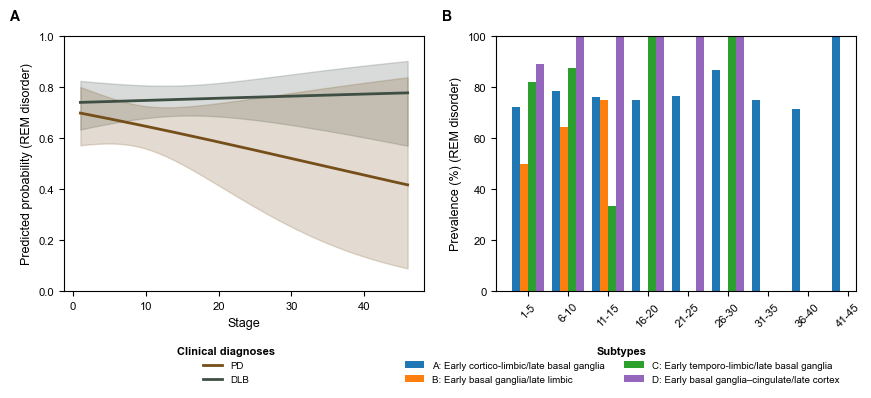

**Supplementary Figure S3: RBD prevalence by SuStaIn stage. A:** Predicted probability of RBD versus SuStaIn stage by clinical diagnosis (PD, DLB) with 95% confidence ribbons from logistic models. **B:** Stage-binned RBD prevalence (%) by subtype (A–D) across the combined cohort. Subtypes: Blue = Subtype A (Early cortico-limbic/late basal ganglia), Orange = Subtype B (Early basal ganglia/late limbic), Green = Subtype C (Early temporo-limbic/late basal ganglia), Purple = Subtype D (Early basal ganglia-cingulate/late cortex). Clinical Diagnoses: Brown = Parkinson’s Disease, Black = Dementia with Lewy Bodies.

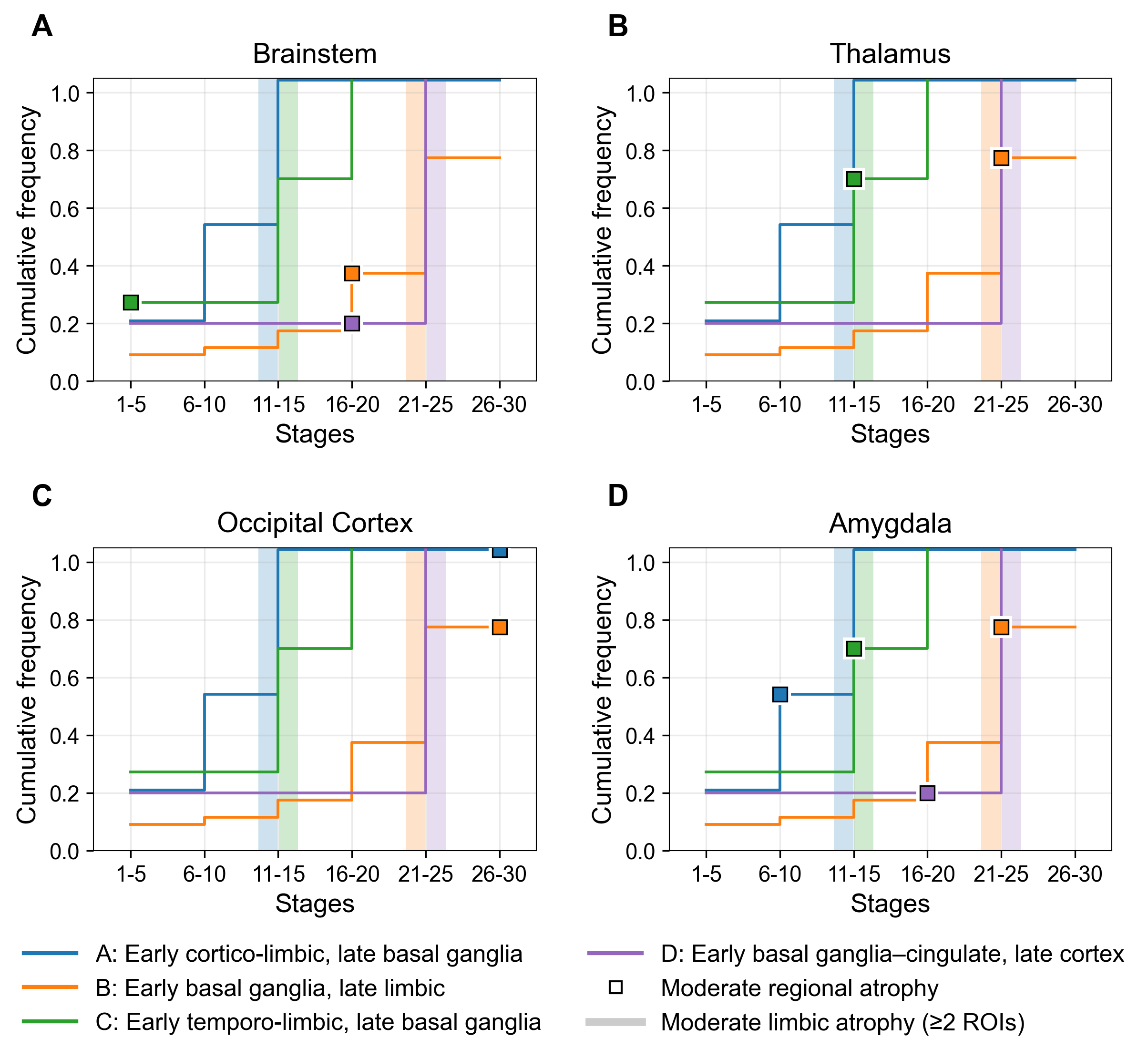

**Supplementary Figure S4: Regional atrophy and stage-dependent emergence of visual hallucinations across subtypes.** Cumulative prevalence of visual hallucinations is plotted against SuStaIn stage for each subtype, alongside the timing of regional atrophy (square markers). Panels show brainstem, thalamus, occipital cortex, and amygdala involvement. The x-axis represents SuStaIn model stage, and the y-axis represents cumulative visual hallucination prevalence. Vertical lines indicate the subtype-specific stage at which the limbic involvement threshold is reached, defined as moderate atrophy, z > 1, in at least two of the four modelled limbic ROIs. Across subtypes, brainstem atrophy often occurred early without a consistent temporal relationship to the rise in visual hallucinations, while thalamic and occipital involvement generally emerged only after a substantial increase in visual hallucination prevalence. In contrast, amygdala involvement aligned more closely with the visual hallucination inflection point in multiple subtypes, supporting a stage-dependent association.

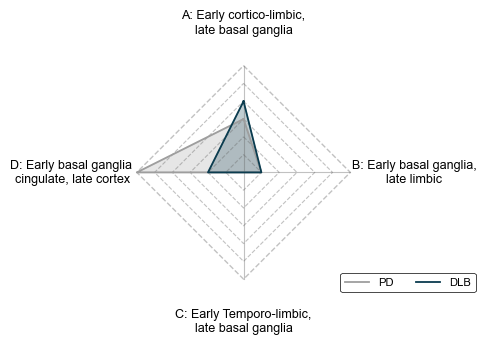

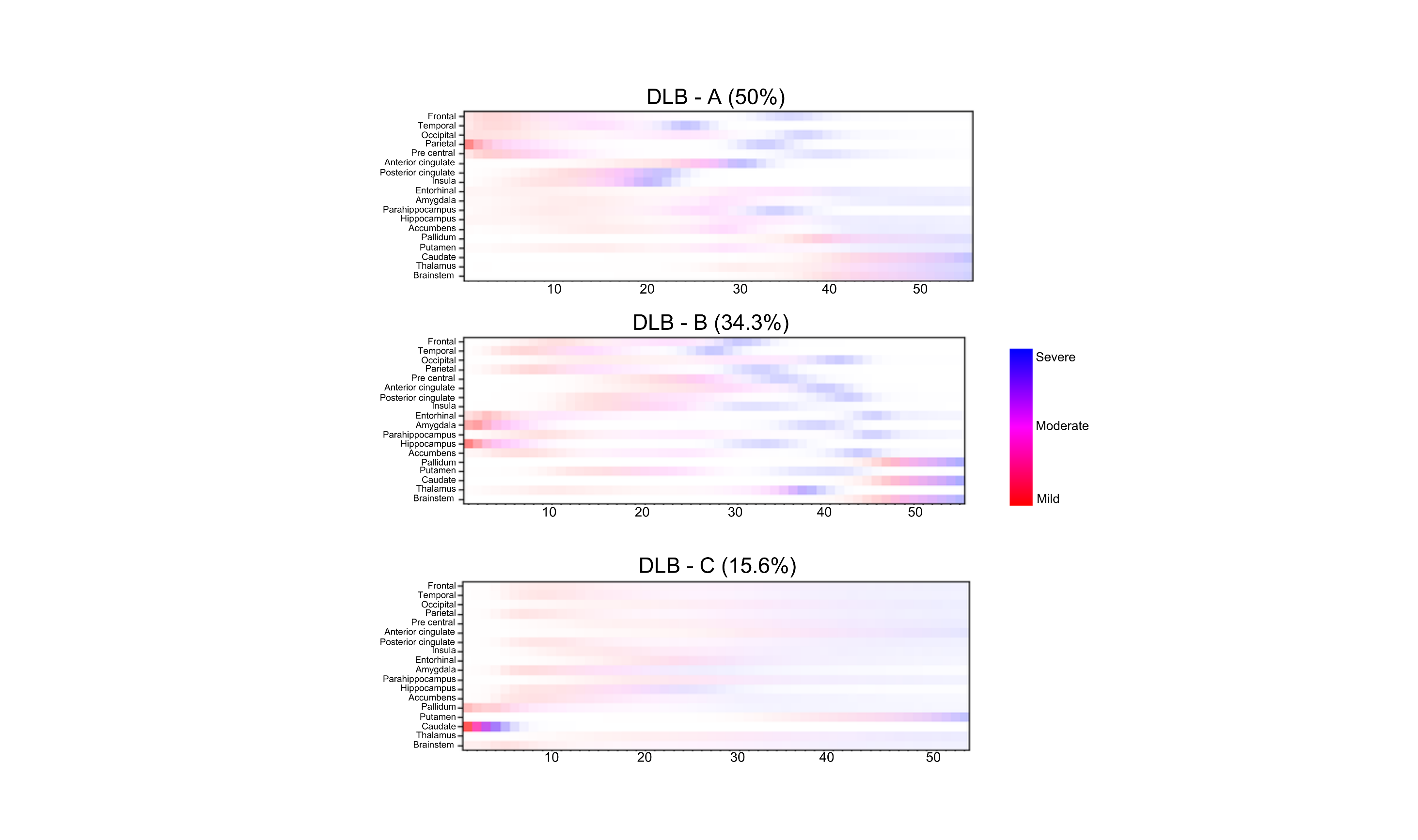

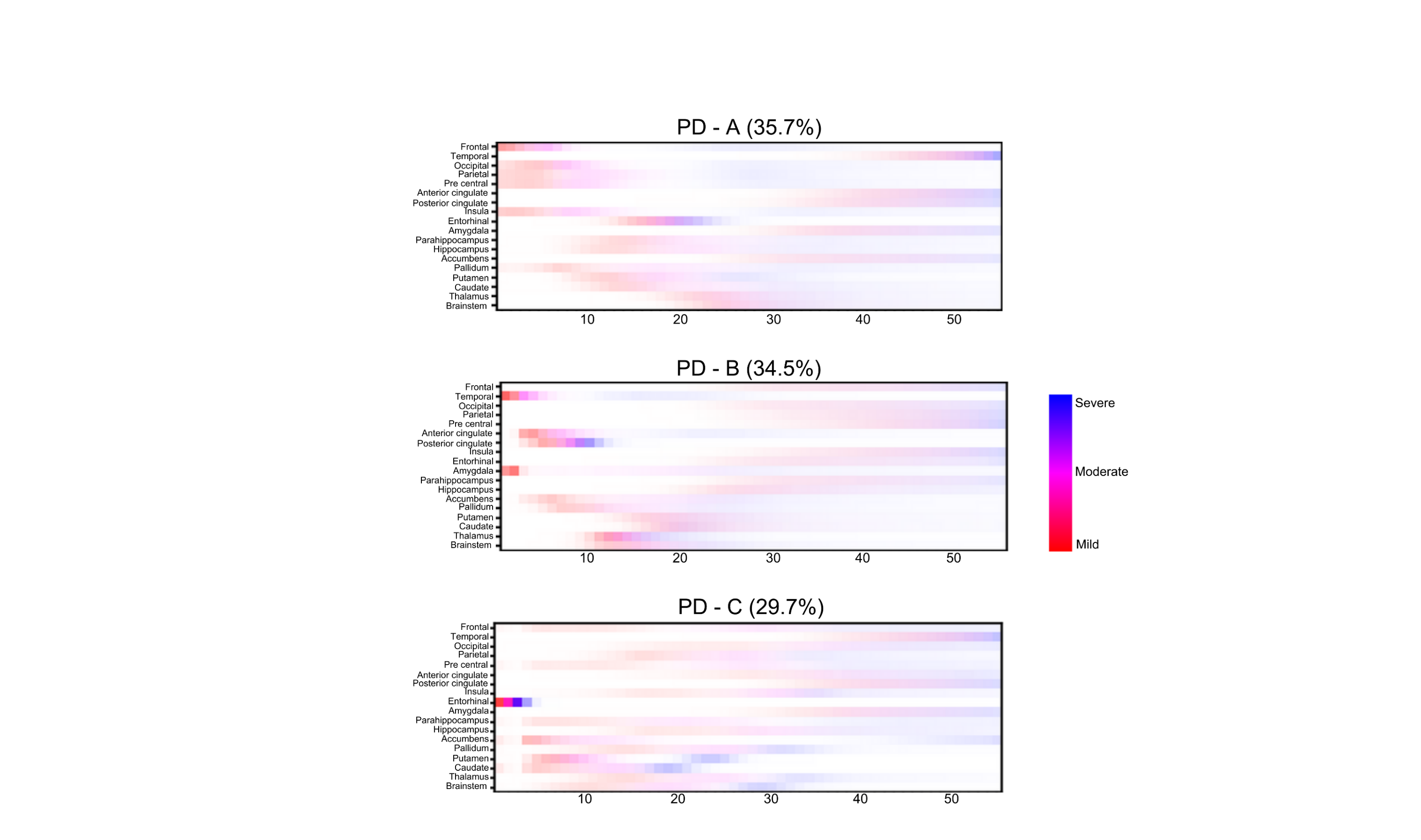

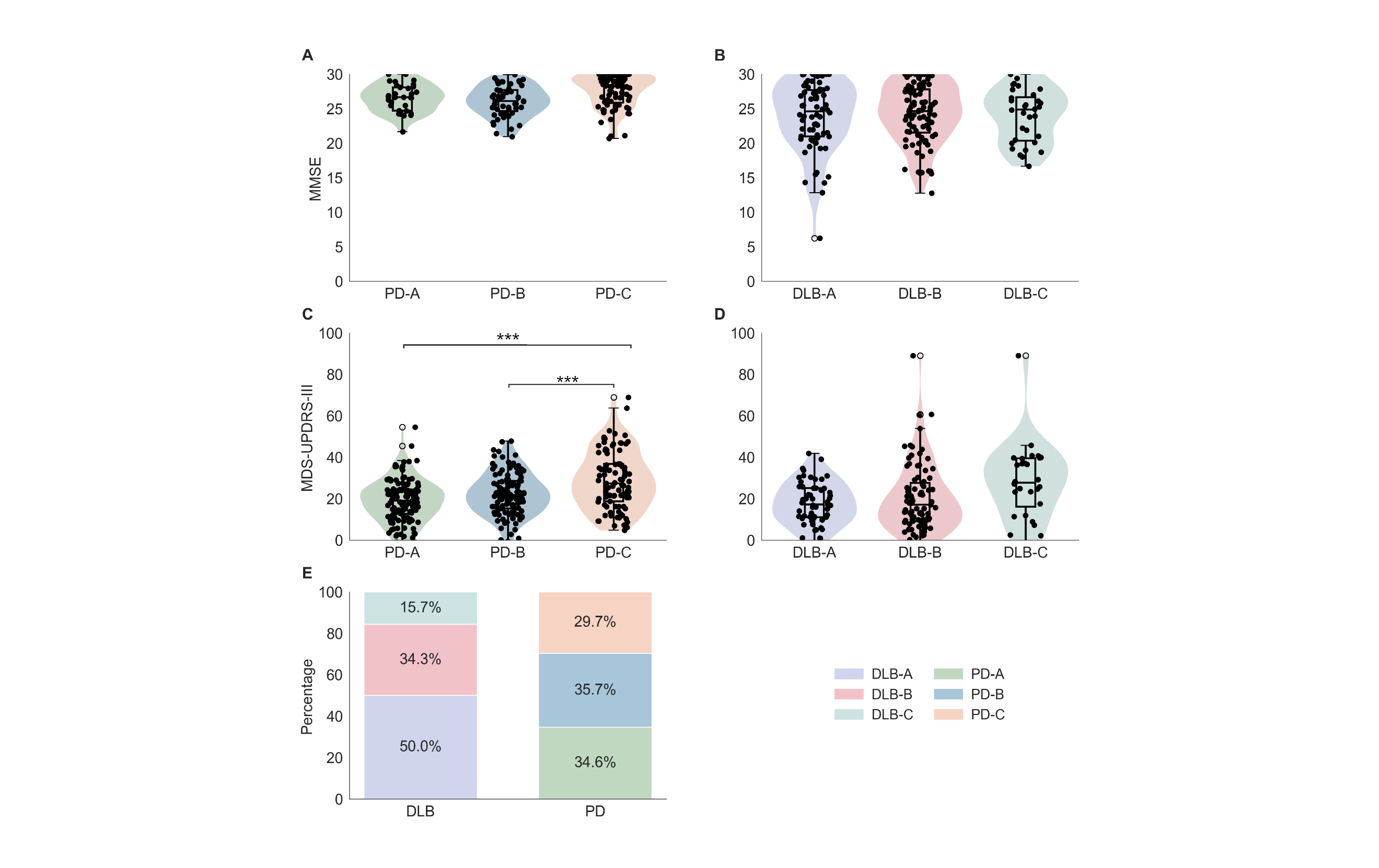

**Supplementary Figure S5: Baseline** subtype in iRBD individuals predicts phenoconversion. Radar plot showing the number of patients with PD (grey) and DLB (dark blue) assigned to each of the four SuStaIn subtypes. The dashed concentric diamonds represent patient counts from 1 (innermost) to 5 (outermost). Blue denotes DLB conversion; grey denotes PD conversion. iRBD to DLB converters concentrate in subtype A, whereas PD shows broader distribution with greater representation in subtype D. PD, Parkinson’s Disease, DLB, dementia with Lewy bodies, iRBD, idiopathic REM sleep behaviour disorder.

**Supplementary Figure S6: Spatiotemporal atrophy signatures of DLB subtypes.** Data-driven subtypes show distinct sequences of regional degeneration in DLB. Positional variance diagrams show atrophy by SuStaIn stage (x-axis) for cortical and subcortical regions (y-axis). Percentages indicate the proportion of the DLB patients in each subtype after excluding stage 0 individuals. Colours indicate severity of each stage, Red: Mild atrophy (z > 0.5), Magenta: Moderate atrophy (z > 1.0), Blue: Severe atrophy (z > 2.0).

**Supplementary Figure S7: Spatiotemporal atrophy signatures of PD subtypes.** Data-driven subtypes show distinct sequences of regional degeneration in PD. Positional variance diagrams show atrophy by SuStaIn stage (x-axis) for cortical and subcortical regions (y-axis). Percentages indicate the proportion of the PD patients in each subtype after excluding stage 0 individuals. Colours indicate severity of each stage, Red: Mild atrophy (z > 0.5), Magenta: Moderate atrophy (z > 1.0), Blue: Severe atrophy (z > 2.0).

**Supplementary Figure S8: Clinical profiles by disease specific subtypes in PD and DLB.** Disease-specific models show limited cognitive and motor differences between subtypes. **A–B,** MMSE by subtype in PD (**A**) and DLB. **C–D:** MDS-UPDRS III by subtype in PD and DLB; asterisks indicate significant contrasts (******* p < 0.001). **E:** Subtype composition (%) within DLB and PD (stacked bars). MMSE, Mini-Mental State Examination; MDS-UPDRS III, Movement Disorder Society Unified Parkinson’s Disease Rating Scale part III; PD, Parkinson’s disease; DLB, dementia with Lewy bodies.
